# *APOE* Genotype and Statin Response: Evidence from Electronic Health Records in the UK Biobank and All of Us Research Program

**DOI:** 10.1101/2024.12.13.24318985

**Authors:** Innocent G. Asiimwe, Andrea L. Jorgensen, Munir Pirmohamed, Mechanism and Therapeutic Research Collaborative

**Author notes:** Correspondence:Innocent G. Asiimwe. Department of Pharmacology and Therapeutics, Institute of Systems, Molecular and Integrative Biology, University of Liverpool, Liverpool, UK., Munir Pirmohamed. Department of Pharmacology and Therapeutics, Institute of Systems, Molecular and Integrative Biology, University of Liverpool, Liverpool, UK.

## Abstract

**Introduction:** APOE genotype may affect statin response. We investigated the relationship between *APOE* genotype and key outcomes in statin users using UK Biobank (UKB) and All of Us (AoU) data.

**Methods:** We analysed electronic health records from up to 45,515 UKB participants and 35,562 AoU participants. Using multivariable linear regression and Cox proportional hazards models, we assessed associations between *APOE* genotype and outcomes, including lipid biomarkers, all-cause mortality, cardiovascular mortality, and major adverse cardiovascular events (MACE).

**Results:** After Bonferroni correction, significant changes in HDLC and triglyceride levels were observed in both cohorts (*P* < 0.01) following statin initiation. For all-cause mortality, significant associations were found in the UKB cohort, with *ε3ε4* (HR: 1.08, 95% CI: 1.01–1.15) and *ε4ε4* (HR: 1.54, 1.33–1.78) carriers showing higher risk compared to the reference *ε3ε3* genotype. In the AoU cohort, only *ε4ε4* carriers showed an increased risk (HR: 1.64, 1.08–2.49). Cardiovascular-related mortality was assessed in only the UKB cohort, with *ε4ε4* carriers having an increased risk (HR: 1.30, 1.01–1.68). In the AoU cohort, lipid level changes were significantly associated with reduced all-cause mortality risk: HDLC (median increase of 0.03 mmol/L, HR: 0.26 [0.16–0.41] per mmol/L), LDLC (median reduction of 0.82 mmol/L, HR: 0.82 [0.69–0.97] per mmol/L), and triglycerides (median reduction of 0.10 mmol/L, HR: 0.79 [0.72–0.87] per mmol/L). No significant associations with MACE were observed in either cohort.

**Conclusion:** This study re-affirms that *APOE* genotype significantly impacts statin response, highlighting the need to integrate genetics into personalized treatment regimens.

## Introduction

Cardiovascular diseases (CVD) are the leading cause of death worldwide.^1^ Statins (HMG-CoA reductase inhibitors) have been shown to reduce CVD-related mortality and are widely used for both primary and secondary prevention.^2^ Apolipoprotein E (*APOE*) genotype has been reported to influence both cardiovascular risk and response to statin treatment.^3–5^ Mutations in two SNPs (rs429358 and rs7412) create three main isoforms: *ε2* (mainly protective), *ε3* (wild-type), and *ε4* (risk-associated). The *ε2* isoform reduces cholesterol levels through lower low-density lipoprotein (LDL) receptor binding, slower lipid clearance, and increased HMG-CoA and LDL receptor synthesis, while *ε4* has the opposite effects.^6–8^ However, carrying two *ε2* copies may impair lipid metabolism (familial dysbetalipoproteinemia), raising CVD risk.^3, 9^

In related work, we conducted a meta-analysis of 52 studies on *APOE* carrier status and statin response.^10^ LDL cholesterol (LDLC), the biomarker most relevant to statin efficacy,^11, 12^ showed greater reductions in *ε2* carriers compared to *ε3* carriers, while *ε4* carriers had a reduced statin response.^10^ Due to limitations such as unknown study quality, high heterogeneity, and few studies focusing on clinical outcomes like mortality, we extended our analysis to the UK Biobank baseline assessment data, with linked mortality records (389,843 to 452,189 participants).^13^ We found that *APOE* genotype was significantly associated with both all-cause death and cardiovascular-related death, with *ε4ε4* carriers having the highest risk. However, the interaction between statin use and *APOE* genotype was not significant. Population-level trends in lipid biomarkers did not always match individual outcomes; while statin-treated *ε4ε4* carriers showed the highest LDLC reductions, these reductions were still lower than those seen in untreated *ε2ε2* individuals. Additionally, the cross-sectional nature of the lipid biomarker analysis limited our ability to track temporal trends, making it difficult to distinguish between individuals with statin-induced decreases and those who had low biomarker levels before statin initiation.^13^ Finally, the UK Biobank baseline dataset lacked relevant details, such as statin doses, as these were not recorded during the verbal interview.^14^

To address the above limitations, particularly to capture individual-level temporal relationships, adjust for statin dose and type, and analyse additional outcomes like major adverse cardiovascular events (MACE), linked electronic health records are required. These records facilitate patient follow-up from pre-statin initiation through post-initiation and, ultimately, to the outcome of interest. Using both a relatively homogeneous dataset (UK Biobank)^14, 15^ and a diverse dataset (All of Us Research Program)^16, 17^ with linked records, we aimed to investigate the relationship between *APOE* genotype and outcomes such as lipid level changes, all-cause mortality, cardiovascular mortality, and MACE in new statin users.

### Methods

The reporting of this study adheres to the REporting of studies Conducted using Observational Routinely collected health Data (RECORD) statement^18^ (Table S1) and the STrengthening the Reporting Of Pharmacogenetic Studies (STROPS) guideline^19^ (Table S2).

### Data Sources

We used two cohorts for our analyses. The first, the UK Biobank (UKB), is a population-based prospective cohort that recruited over half a million participants from 22 assessment centres across England, Scotland, and Wales between 2006 and 2010.^14, 15^ Approximately 230,000 participants have linked healthcare records, including death and cancer registries, hospital inpatient and outpatient episodes, and primary care data. Ethical approval was received by the UKB (North-West Multicentre Research Ethics Committee approval number: 11/NW/0382), and written informed consent was received from all participants. Our study was approved by the UKB (application number: 56653).

We also used data from the All of Us (AoU) Research Program, launched by the US National Institutes of Health in 2018 to enhance diversity in biomedical research.^16^ To date, the program has enrolled over 700,000 participants, with 80% from historically underrepresented backgrounds. Its Dataset v7 repository includes electronic health records (EHR) and genomic data, accessible through a cloud-based Researcher Workbench. Access to genomic data requires Controlled tier access, granted after completing registration, training, and a data-use agreement.^16, 17^

### Study Participants

We included only participants with genomic data who met genotype quality control criteria: a) consistent genetic and self-identified sex, b) no sex chromosome aneuploidy, c) non-outliers for heterozygosity or missing rate, and d) unrelated to other participants, as described elsewhere.^13^ Additional inclusion criteria were: a) prescription of one of the following statins: atorvastatin, cerivastatin, fluvastatin, lovastatin, pitavastatin, pravastatin, rosuvastatin, or simvastatin (note: lovastatin and pitavastatin are not available in the UK, and cerivastatin was withdrawn shortly after approval),^20^ b) more than one year of EHR data, c) no statin prescription within the first year of registration (UKB) or first interaction with the healthcare system (AoU, defined as the first record in any of these Observational Medical Outcomes Partnership [OMOP] tables: ’Condition Occurrence,’ ’Device Exposure,’ ’Drug Exposure,’ ’Measurement,’ ’Observation,’ ’Observation Period,’ ’Procedure Occurrence,’ or ’Visit Occurrence’), ensuring inclusion of only incident users, d) statin treatment for at least three months (84 days), and, for lipid outcome analyses, e) at least two biomarker measurements – one before starting statins and one at least 28 days after. In a sensitivity analysis, as in the UK Biobank baseline analysis,^13^ we also excluded individuals with dysbetalipoproteinemia (*ε2ε2* homozygotes with pre-treatment total cholesterol ≥ 200 mg/dL [5.2 mmol/L] and triglycerides ≥ 175 mg/dL [2.0 mmol/L]).^21, 22^

To identify statin users, in UKB, we used Resource 592 (“Clinical coding classification systems and maps,” https://biobank.ndph.ox.ac.uk/ukb/refer.cgi?id=592) to match statin generic names to British National Formulary brand names, which were then mapped to prescription records. In AoU, we used the Anatomical Therapeutic Chemical (ATC) Classification System code ’C10AA’ (HMG CoA reductase inhibitors) along with curated phenotyping codes from the “All by All - Drug Phenotypes Curation” Workbench (https://workbench.researchallofus.org/workspaces/aou-rw-046fb18c/allbyalldrugphenotypescuration/data).

### Outcome and Follow-up

For lipid biomarker analysis, we included those available for at least 781 eligible participants (see "Sample size and Power analysis" section): four biomarkers from UKB (all in mmol/L: HDL Cholesterol [HDLC], LDL Cholesterol [LDLC], Total Cholesterol [TC], and Triglycerides [TG]) and five from AoU (all in mg/dL: HDLC, non-HDLC, LDLC, TC, and TG). For consistency, the AoU units (mg/dL) were converted to mmol/L using conversion factors of 88.57 for triglycerides and 38.67 for other lipid biomarkers. In the UKB, we identified biomarker measurements using Read v2 codes from the Health Data Research (HDR) UK Phenotype Library (https://phenotypes.healthdatagateway.org/phenotypes/) and a previous publication.^23^ To ensure data quality, we excluded outliers, defined as values outside 1.5 times the interquartile range. For AoU, we used curated phenotyping codes from the “All by All - Lab Measurements Phenotypes Curation” Workbench (https://workbench.researchallofus.org/workspaces/aou-rw-0c74a4d9/allbyalllabmeasurementsphenotypescuration/data), harmonizing units, excluding outliers (boundaries set by clinical experts), and excluding data from sites with quality concerns post-harmonization. In addition to analysing net biomarker changes, we calculated percentage changes using the formula: percentage change = ((median biomarker value four weeks post-treatment – median biomarker value pre-treatment) / median biomarker value pre-treatment) * 100. Post-treatment values were taken as the median measurement starting four weeks after treatment initiation, allowing adequate time for statins to take effect. We tested various timeframes for capturing biomarker values: a) no time limit (primary analysis with all values), b) one year before and after treatment, c) six months before and one year after treatment, and d) six months before and after treatment.

The clinical outcomes assessed included all-cause mortality, cardiovascular-related deaths, and major adverse cardiovascular events (MACE). Cardiovascular deaths were identified using ICD-10 codes (I10–15, I44–51, I20–25, I61–73) following the European Systematic Coronary Risk Evaluation (SCORE) guidelines.^24, 25^ MACE was defined per the Ramirez et al. criteria, which included ICD-9 and ICD-10 codes for ischemic heart disease, myocardial infarction, heart failure, and ventricular arrhythmia.^26^ In the UKB, MACE cases were identified through Category 1712 ("Health-related outcomes first occurrences", https://biobank.ndph.ox.ac.uk/ukb/label.cgi?id=1712) from primary care data (Category 3000), hospital inpatient data (Category 2000), and death registry records (Fields 40001 and 40002). For AoU, MACE cases were identified from ’Condition Occurrence,’ ’Observation,’ ’Measurement,’ and ’Procedure Occurrence’ OMOP tables, with death data sourced from the ’Death’ tables. However, over 98% of deaths in AoU lacked cause information or matching concepts in the death table, preventing identification of cardiovascular-related deaths in the AoU data.

We assumed continuous statin exposure from first statin prescription to the end of prescription records. Participants were followed from their first statin prescription until the earliest of the outcome occurrence (death or MACE) or the censor date (UKB: 31^st^ December 2022 for mortality outcomes and 31^st^ May 2016 for MACE [based on the earliest primary care censoring date, https://biobank.ndph.ox.ac.uk/ukb/exinfo.cgi?src=Data_providers_and_dates]; AoU: 1^st^ July 2022).

### Predictors

The exposure variable was *APOE* genotype (*ε2ε2*, *ε2ε3*, *ε2ε4*, *ε3ε3*, *ε3ε4* and *ε4ε4*, determined based on the SNPs rs429358 and rs7412),^27^ and statin use. Genotyping, imputation, and quality control procedures for both UKB and AoU have been described previously.^15, 17^

In the UKB baseline analysis,^13^ we adjusted for several baseline covariates, including education level, socioeconomic status, body mass index, alcohol and smoking habits, and self-reported physical activity.

However, these covariates are not stable over time and may differ between the date of UKB recruitment and the date of the first statin prescription. Indeed, only 15.6%, 28.9%, and 66.0% of analysed UKB participants had measurements for these covariates within 1, 2, and 5 years of the index date (start of follow-up), respectively (see Figure S1). Given the potential changes by the index date, we limited our analysis to more stable factors: age at follow-up start, sex, race, genotyping array (UKB only), the first ten principal components of genetic ancestry, and comorbidities, including diabetes (ICD-10: E10-E14), hypertension (ICD-10: I10-I15), and cardiovascular disease (ICD-10: I20–25, I60–64, per American College of Cardiology/American Heart Association guidelines,^24, 28^ and MACE-related codes^26^). We also included data provider (UKB only: Vision England, TPP England, Vision/EMIS Health Scotland, Vision/EMIS Health Wales, as AoU had over 45 sites for eligible participants), statin type, and statin strength as covariates.

Lastly, net changes in lipid biomarkers were added as covariates to evaluate their association with all-cause mortality. For HDLC, net changes were calculated as post-treatment minus pre-treatment levels, with HRs reflecting the risk associated with a 1 mmol/L increase (HR < 1 indicates a beneficial effect). For other biomarkers, net changes were calculated as pre-treatment minus post-treatment levels, with HRs representing the risk associated with a 1 mmol/L reduction (HR < 1 indicates a beneficial effect).

### Sample size and Power analysis

We aimed to analyse all eligible participants in the available cohorts to maximize sample size, so we report statistical power rather than minimum sample size requirements for clinical outcomes. For lipid biomarkers, which had varying sample sizes, we calculated the minimum sample size needed for inclusion.

For lipid biomarker analysis (using percent change as the outcome), we determined the sample size with the ss.calc.linear function from the R package "genpwr".^29^ Assuming a target coefficient of determination (*R²*) of 1%, minor allele frequencies of 5–20%, 80% power, standard deviations of 10–30%, and an additive mode of inheritance, a minimum of 781 participants was required at *P* = 0.05. After applying Bonferroni correction for 2, 3, 4, and 5 biomarkers, the minimum sample sizes increased to 946, 1042, 1110, and 1163, respectively.

For clinical outcomes, we used the powerEpiInt function from the R package powerSurvEpi^30^ to estimate power, requiring a pilot dataset and two binary variables. We included sex (in addition to *APOE* genotype) due to its known influence on *APOE* effects in statin-treated patients.^10^ For each genotype contrast versus *ε3ε3*, we randomly selected 1,000 participants, with assumptions of a 0.05 significance level, effect sizes (HRs) of 1.1–2.3, and an 80% power threshold.

In the UKB, a sample size of 45,512 was adequate to detect all-cause mortality effects across genotypes with minimum detectable HRs ranging from 1.2 (*ε3ε4*) to 2.3 (*ε2ε2*) (Figure S2). For cardiovascular deaths, there was insufficient power for *ε2ε2*, with minimum HRs required for detection of other genotypes ranging from 1.3 to 1.8. For MACE (sample size: 45,146), *ε2ε2* was detectable at an HR of about 1.8, and other genotypes at HRs between 1.1 and 1.3. In AoU, a sample size of 35,562 was sufficient for detecting all-cause mortality effects only for the genotypes *ε2ε3* (HR = 2.3) and *ε3ε4* (HR > 1.5). For MACE, all genotypes were detectable, requiring HRs ranging from 1.1 to 1.8 (Figure S2).

### Missing Data

Participants with missing outcome data were excluded from the corresponding analyses. The UKB dataset had 0.02% participants missing statin strength data and these were excluded. On the other hand, the AoU dataset had missing strength data for 14.7% of participants. Assuming statin strength was missing at random, we imputed these missing values using Multivariate Imputation by Chained Equations (MICE) in R^31^ (Figure S3). The imputation model included all covariates (see ‘Predictors’ section) and outcomes (mortality, mace, pre-treatment biomarker levels, and post-treatment biomarker levels),^32^ with predictive mean matching as the imputation method. We generated 20 imputed datasets, exceeding the percentage of incomplete cases,^33^ and pooled estimates from the imputed datasets using Rubin’s rules.^34^ In a sensitivity analysis, we analysed only the participants with complete data.

### Statistical Analysis

For lipid outcomes (net or percentage changes in biomarker levels), we used linear regression models, and for mortality outcomes, we applied Cox proportional hazard models with competing risks (two terminal states),^35^ such as all-cause death for cardiovascular death or MACE, accounted for. When participants were diagnosed with MACE on the same day as their recorded death, they were classified as MACE cases, as death cannot occur before a MACE event. Each model adjusted for the covariates listed in the ’Predictors’ section, with certain categorical factors combined due to low sample sizes. Specifically, statins were categorized into ’Simvastatin,’ ’Atorvastatin,’ and ’Other’, as the remaining statin types had low frequencies. Effects were reported as regression coefficients for linear regression and hazard ratios (HRs) with 95% confidence intervals (CIs) for Cox regression. To generate trend p-values for *APOE* genotype, we used Type III ANOVA tests from the car package.^36^ Statistical significance was set at *P* = 0.05, with Bonferroni correction for multiple testing. All analyses were conducted in R (version 4.4.1).^37^

### Patient and public involvement

Patients and their representatives were not involved in formulating the research question or selecting outcomes for this analysis. However, they are actively engaged in both the UKB and the AoU Research Program.

## Results

### Participants

From half a million UK Biobank (UKB) participants, we selected 200,672 unrelated individuals with quality-controlled *APOE* data and linked primary care records (Figure 1). After exclusions based on statin use and registration record length, our analysis cohort for mortality outcomes included 45,515 incident statin users. Biomarker analyses were conducted on specific subsets: HDL cholesterol (HDLC: 5,736), LDL cholesterol (LDLC: 5,052), total cholesterol (TC: 6,560), and triglycerides (TG: 5,462). MACE analyses included 45,149 participants. Using similar eligibility criteria in the All of Us (AoU) cohort, from 189,478 unrelated participants with electronic health records and quality-controlled *APOE* data, we identified 35,562 incident statin users for mortality and MACE outcomes, with biomarker analyses on subsets: HDLC (17,626), non-HDLC (4,931), LDLC (11,235), TC (18,484), and TG (17,712).

**Figure 1.**
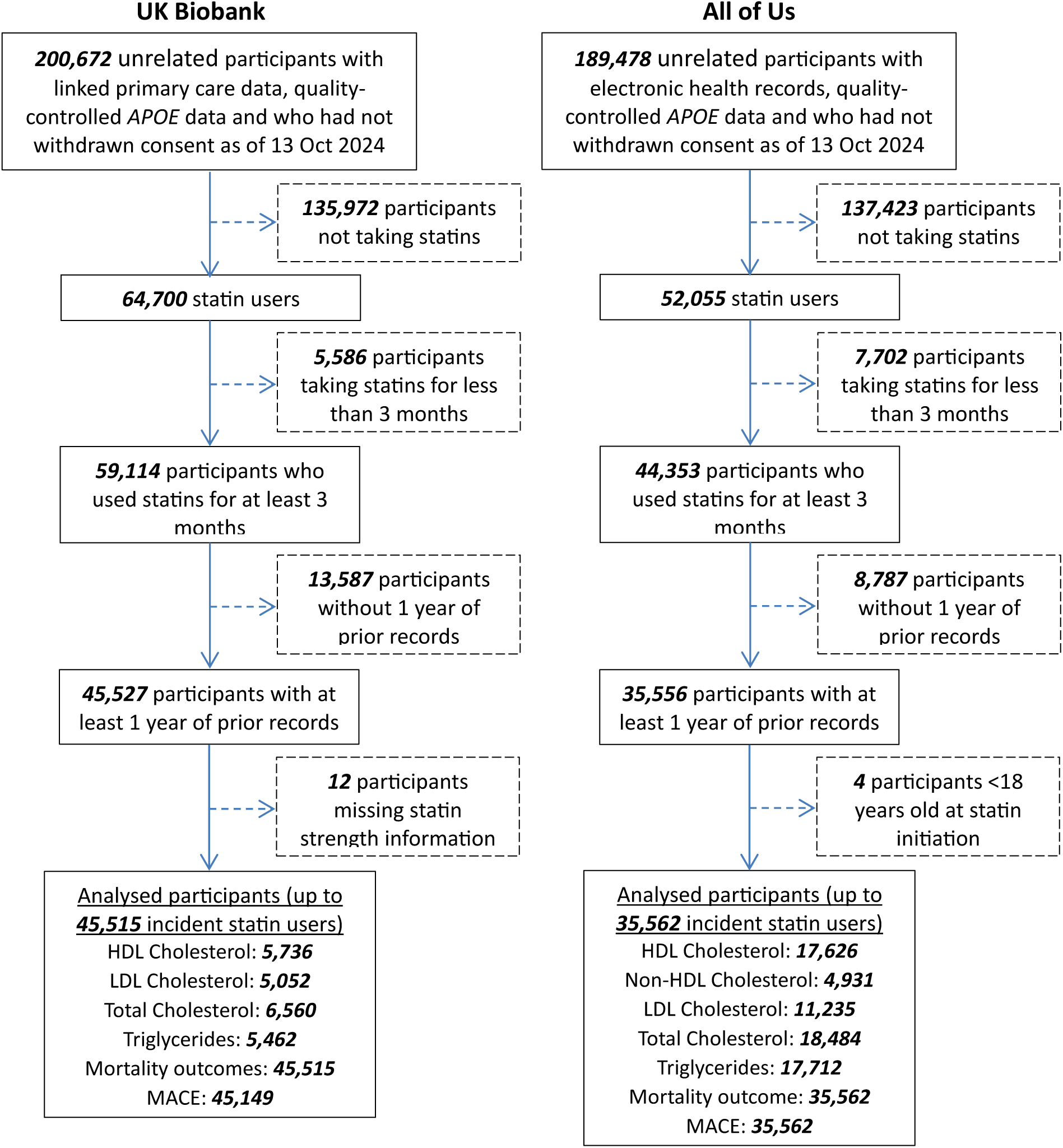
Flow chart for included participants. Bold values represent the total number of participants at each stage. *APOE* = Apolipoprotein E, HDL = high-density lipoprotein, LDL = low-density lipoprotein, MACE = major adverse cardiovascular events.

Table 1 shows the characteristics of included participants. The median age in both cohorts was around 60 years (UKB: 60.3 years; AoU: 59.1 years). The UKB cohort had a lower proportion of females (41.9% vs.

**Table 1.** Characteristics of UK Biobank and All of Us program participants.

|  | UK Biobank (N=45515) | All of Us Program (N=35562) |
| --- | --- | --- |
| **Age (years)** |  |  |
| Mean (SD) | 60.3 (7.34) | 59.1 (10.9) |
| Median [Min, Max] | 60.9 [26.7, 78.8] | 59.8 [18.2, 98.0] |
| **Sex** |  |  |
| Female | 19092 (41.9%) | 19123 (53.8%) |
| Male | 26423 (58.1%) | 16439 (46.2%) |
| **Race** |  |  |
| White | 42915 (94.3%) | 24548 (69.0%) |
| Asian | 1496 (3.3%) | 702 (2.0%) |
| Black | 413 (0.9%) | 6734 (18.9%) |
| Mixed/Other/Unknown | 691 (1.5%) | 3578 (10.1%) |
| **Hypertension history** |  |  |
| No | 17320 (38.1%) | 10660 (30.0%) |
| Yes | 28195 (61.9%) | 24902 (70.0%) |
| **Diabetes Mellitus history** |  |  |
| No | 35860 (78.8%) | 24245 (68.2%) |
| Yes | 9655 (21.2%) | 11317 (31.8%) |
| **Cardiovascular disease history** |  |  |
| No | 27377 (60.1%) | 27500 (77.3%) |
| Yes | 18138 (39.9%) | 8062 (22.7%) |
| **With dysbetalipoproteinemia** |  |  |
| No | 45354 (99.6%) | 35438 (99.7%) |
| Yes | 161 (0.4%) | 124 (0.3%) |
| **Data provider** |  |  |
| England TPP | 35421 (77.8%) | - |
| England Vision | 4525 (9.9%) | - |
| Scotland | 5451 (12.0%) | - |
| Wales | 118 (0.3%) | - |
| **Statin type** |  |  |
| Atorvastatin | 10410 (22.9%) | 17883 (50.3%) |
| Cerivastatin | 144 (0.3%) | 9 (0.0%) |
| Fluvastatin | 257 (0.6%) | 1893 (5.3%) |
| Lovastatin | - | 1019 (2.9%) |
| Pitavastatin | - | 58 (0.2%) |
| Pravastatin | 1014 (2.2%) | 3127 (8.8%) |
| Rosuvastatin | 583 (1.3%) | 2913 (8.2%) |
| Simvastatin | 33107 (72.7%) | 8660 (24.4%) |
| **Statin strength** |  |  |
| Mean (SD) | 28.9 (14.4) | 28.1 (19.1) |
| Median [Min, Max] | 40.0 [0.100, 80.0] | 20.0 [0.300, 80.0] |
| Missing<sup>a</sup> | - | 5222 (14.7%) |
| **Genotyping array** |  |  |
| UK Biobank Axiom array | 40149 (88.2%) | - |
| UK BiLEVE Axiom array | 5366 (11.8%) | - |
| **APOE genotype** |  |  |
| ε2ε2 | 229 (0.5%) | 231 (0.6%) |
| ε2ε3 | 4436 (9.7%) | 3691 (10.4%) |
| ε2ε4 | 1019 (2.2%) | 784 (2.2%) |
| ε3ε3 | 26963 (59.2%) | 21702 (61.0%) |
| ε3ε4 | 11646 (25.6%) | 8266 (23.2%) |
| ε4ε4 | 1222 (2.7%) | 888 (2.5%) |
| **All-cause death** |  |  |
| No | 40003 (87.9%) | 34842 (98.0%) |
| Yes | 5512 (12.1%) | 720 (2.0%) |
| **Cardiovascular-related death** |  |  |
| No | 43283 (95.1%) | - |
| Yes | 2232 (4.9%) | - |
| **Death follow-up time (years)** |  |  |
| Mean (SD) | 13.6 (4.76) | 6.80 (5.53) |
| Median [Min, Max] | 13.5 [0.282, 33.8] | 5.43 [0.230, 34.2] |
| **MACE** |  |  |
| Censored | 33510 (73.6%) | 25481 (71.7%) |
| Got event | 11639 (25.6%) | 10081 (28.3%) |
| Missing | 366 (0.8%) | - |
| **MACE follow-up time (years)** |  |  |
| Mean (SD) | 5.94 (4.44) | 5.13 (4.99) |
| Median [Min, Max] | 5.29 [0.00274, 26.5] | 3.54 [0.00274, 32.4] |
| Missing | 366 (0.8%) | - |

<sup>a</sup>12 (0.02%) UK Biobank participants missing strength information not included.
**Abbreviations:** APOE = Apolipoprotein E, BiLEVE = Biobank Lung Exome Variant Evaluation, MACE = major adverse cardiovascular events, Max = Maximum, Min = Minimum, N = sample size, SD = standard deviation.

53.8%) and was less racially diverse (94.3% White vs. 69.0% White). Hypertension and diabetes mellitus were less prevalent the UKB cohort (61.9% vs. 70.0% and 21.2% vs 31,8% respectively) while cardiovascular disease history was more frequent in the UKB cohort (39.9% vs. 22.7%). Dysbetalipoproteinemia was rare in both cohorts (0.4% in UKB and 0.3% in AoU). Atorvastatin and simvastatin were the most initially prescribed statins in both cohorts, though their distribution differed: 72.7% of UKB participants were on simvastatin compared to 24.4% in AoU, while 50.3% of AoU participants were on atorvastatin compared to 22.9% in UKB. *APOE* genotypes were similarly distributed, with *ε3ε3* being the most common (UKB: 59.2%; AoU: 61.0%) followed by *ε3ε4* (25.6% vs. 23.2%). The UKB cohort showed a higher all-cause mortality rate (12.1% vs. 2.0%), although fewer participants experienced MACE (25.6% vs. 28.3%). Cardiovascular-related deaths were obtained only for the UKB cohort (4.9%). Median follow-up times were longer in the UKB cohort than in AoU (13.5 years vs. 5.43 years for death follow-up; 5.29 years vs. 3.54 years for MACE follow-up). Additional participant characteristics, including stratifications by mortality and each biomarker, as well as details on the principal components of genetic ancestry and net and percentage changes for each biomarker, are provided in Tables S3 and S4. For example, the median percentage changes in lipid biomarkers were: HDLC (0.0%), LDLC (−35.0%), TC (−22.5%), and TG (−11.8%) in the UKB cohort, and HDLC (2.33%), non-HDLC (−26.2%), LDLC (−26.6%), TC (−16.6%), and TG (−8.16%) in the AoU cohort. Figure S4 shows the correlations between biomarker levels before statin treatment and their net changes after treatment.

### Lipid outcomes

Tables S5 (net change) and S6 (percentage change) summarize associations between all analysed covariates and the lipid biomarkers, while Figure 2 shows the relationship between *APOE* genotype and net and percentage changes in these biomarkers. After applying Bonferroni corrections for four lipid outcomes in UKB (threshold: 0.0125) and five in AoU (threshold: 0.01), only net changes in HDLC (AoU, *P* < 0.001) and TG (*P* < 0.001 in both UKB and AoU) were significant. HDLC in UKB (*P* = 0.020) was slightly above the threshold. To account for baseline differences, percentage changes were analysed, with HDLC (UKB, *P* = 0.006; AoU, *P* = 0.003) and TG (UKB, *P* = 0.002; AoU, *P* < 0.001) reaching significance. Excluding individuals with dysbetalipoproteinemia (0.3–0.4%) yielded similar results (Figure S5), with the main difference being a better alignment in terms of the effect direction for *ε2ε2* homozygotes with other *ε2* carriers, although confidence intervals widened due to smaller sample size.

**Figure 2.**
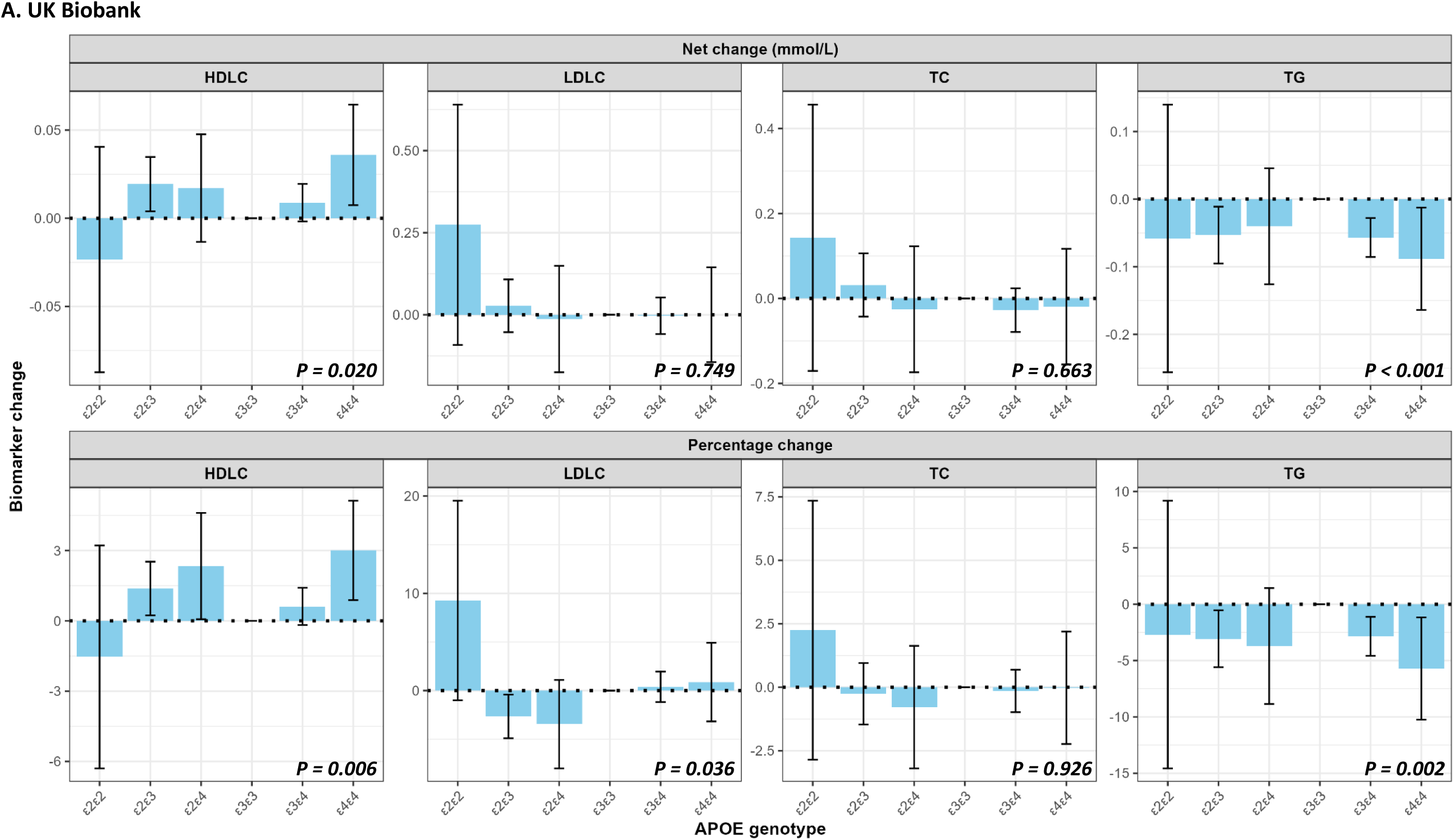
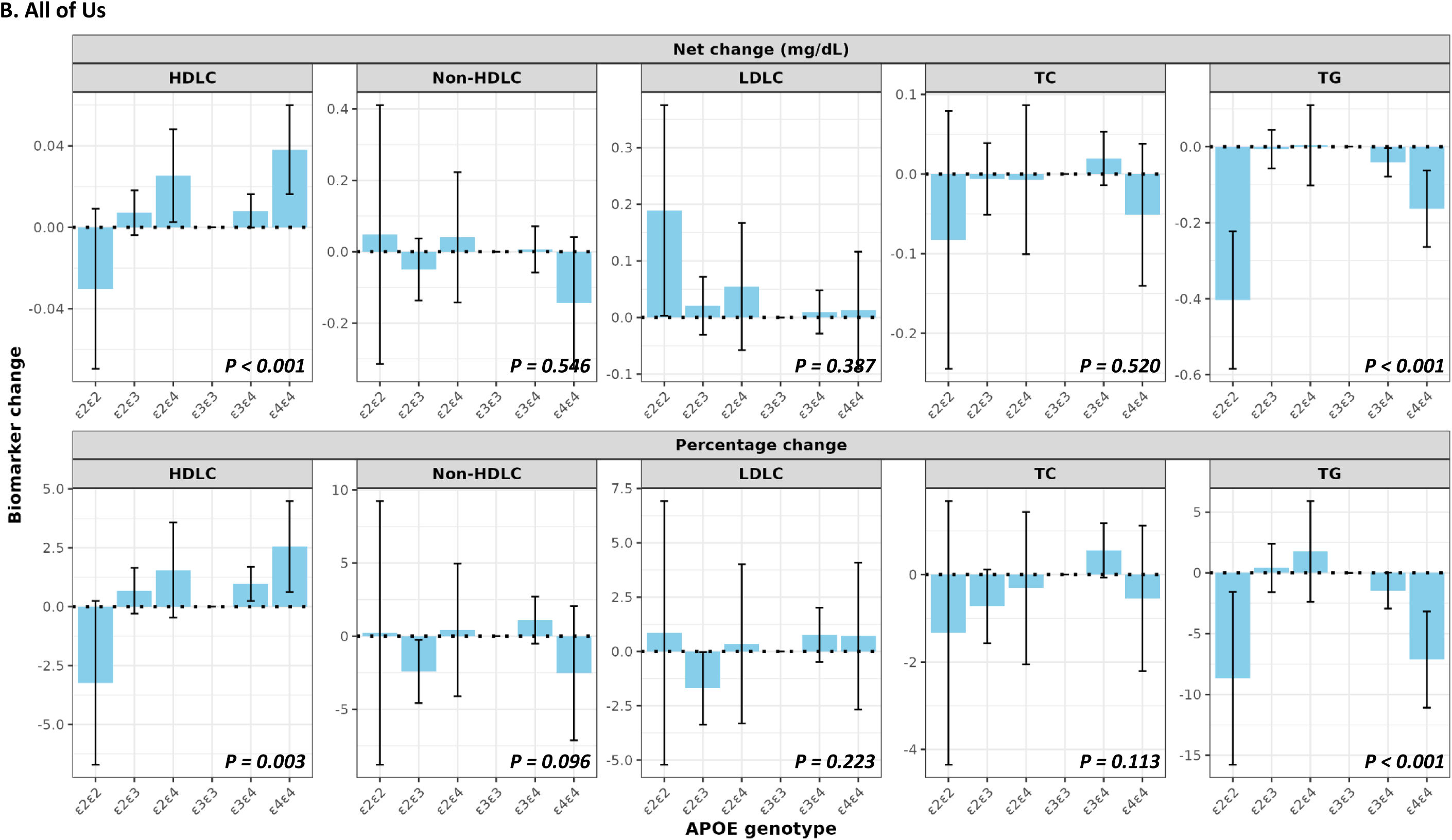
Net and percentage changes in lipid biomarkers stratified by *APOE* genotype. **A.** UK Biobank. **B.** All of Us Program. The top row displays net changes, and the bottom row shows percentage changes. For HDLC (increase beneficial), positive values indicate more benefit with statins relative to the *ε3ε3* genotype, while for other biomarkers (reduction beneficial), negative values indicate more benefit. Error bars represent 95% confidence intervals. *APOE* = Apolipoprotein E, HDLC = high-density lipid cholesterol, LDLC = low-density lipid cholesterol, TC = Total cholesterol, TG = triglycerides.

Initial analyses applied no time limits to maximize sample size, but sensitivity analyses with time restrictions (Figure S6) reduced sample sizes to about 60–75% for one year before and after statin initiation, 50–60% for six months before and one year after, and 30–45% for six months before and after. Figure S7 shows percentage changes in lipid biomarkers by *APOE* genotype across these time windows, with confidence intervals widening as sample sizes decreased.

### Mortality and MACE outcomes

Table S7 presents the associations between all covariates and mortality outcomes, while Figures 3 (UKB) and 4 (AoU) focus specifically on the effects of *APOE* genotype and statin use on these outcomes. In Figure 3, Panel A (UKB cohort), *APOE ε4ε4* carriers had a significantly higher risk of all-cause mortality compared to the reference *ε3ε3* genotype (HR: 1.54, 95% CI: 1.33–1.78), while *ε3ε4* carriers showed a modestly elevated risk (HR: 1.08, 95% CI: 1.01–1.15). In Figure 4, Panel A (AoU cohort), although the trend *P*-value (0.371) was not significant, *ε4ε4* carriers showed a significantly increased risk of all-cause mortality (HR: 1.64, 95% CI: 1.08–2.49), whereas *ε3ε4* carriers showed a non-significant increase (HR: 1.07, 95% CI: 0.89–1.28). Sensitivity analyses, including the exclusion of participants with dysbetalipoproteinemia and complete-case analysis for the AoU cohort, produced results consistent with the primary analysis (Table S7).

**Figure 3.**
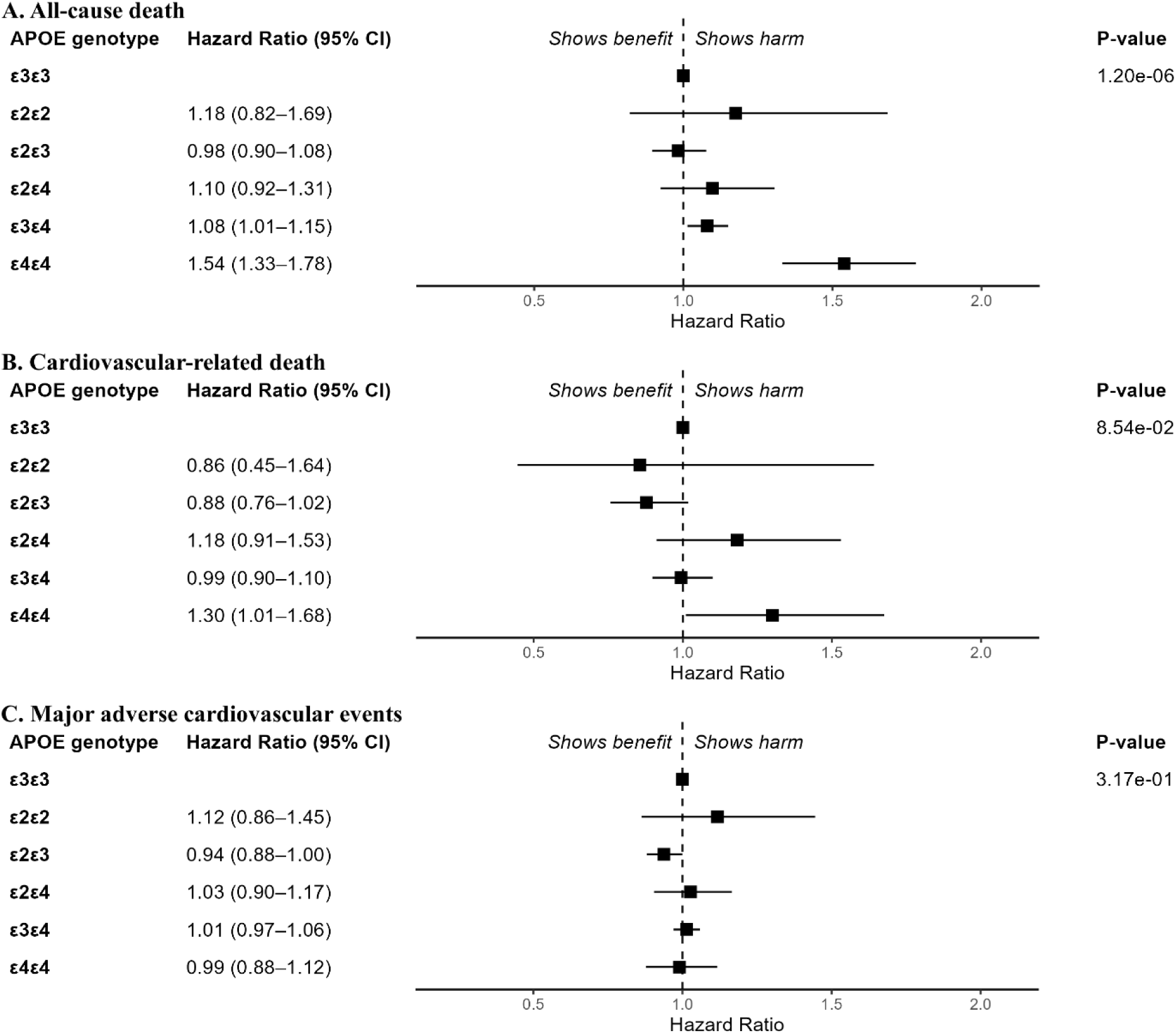
Associations between *APOE* genotype and clinical outcomes in the UK Biobank. This figure shows hazard ratios and 95% confidence intervals for three clinical outcomes: **A.** All-cause death, **B.** Cardiovascular- related death, and **C.** Major adverse cardiovascular events. The analyses are adjusted for the following covariates: age, sex, histories of hypertension, diabetes mellitus, and cardiovascular disease, data provider, type and strength of statin, genotyping array, and the first ten principal components of genetic ancestry. *APOE* = Apolipoprotein E.

**Figure 4.**
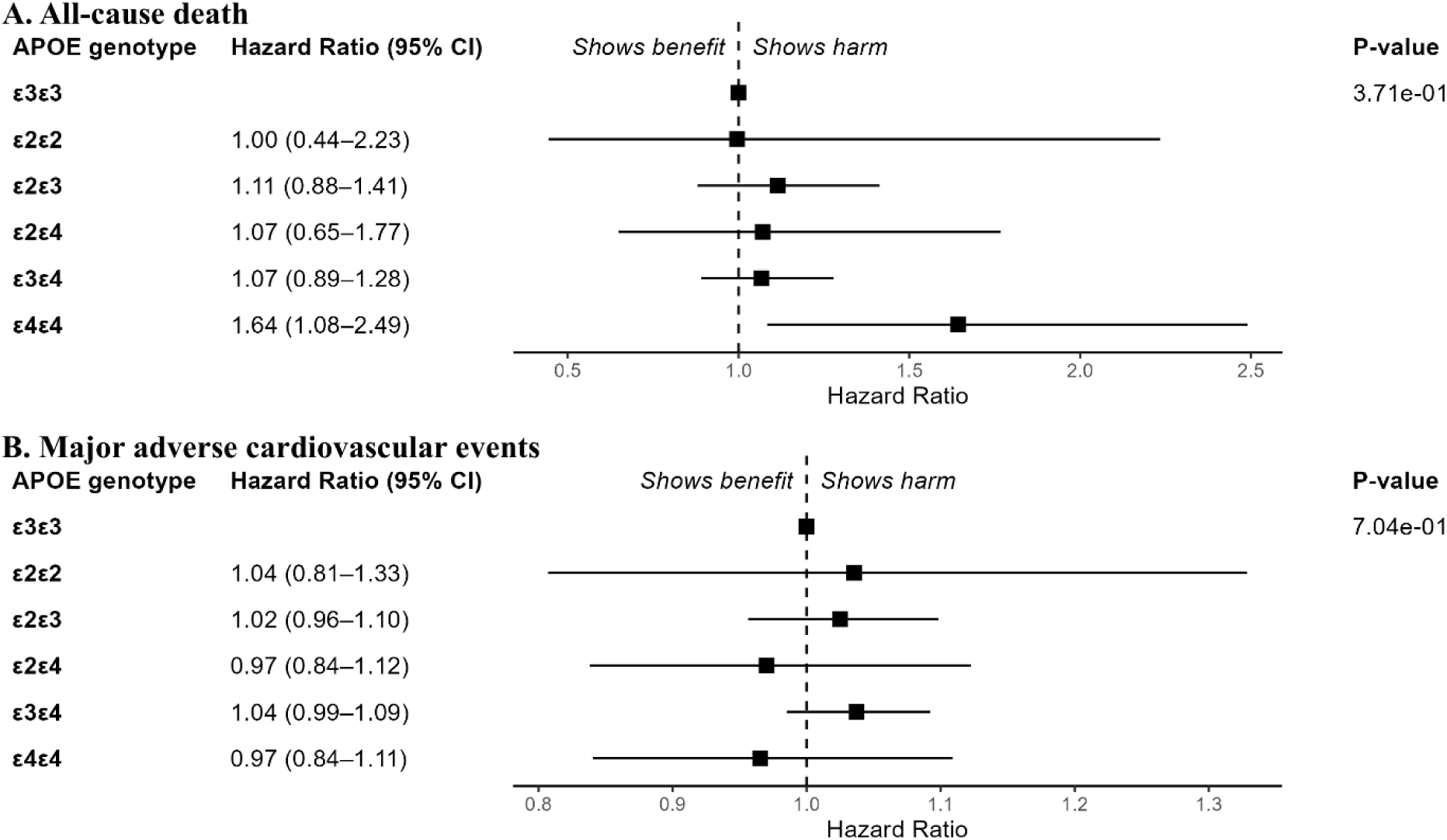
Associations between *APOE* genotype and clinical outcomes in the All of US Program. This figure shows hazard ratios and 95% confidence intervals for three clinical outcomes: **A.** All-cause death and **B.** Major adverse cardiovascular events. The analyses are adjusted for the following covariates: age, sex, histories of hypertension, diabetes mellitus, and cardiovascular disease, type and strength of statin, and the first ten principal components of genetic ancestry. *APOE* = Apolipoprotein E.

Cardiovascular-related mortality was assessed only in the UKB cohort (Figure 3, Panel B) where only *ε4ε4* carriers demonstrated a significantly increased risk (HR: 1.30, 95% CI: 1.01–1.68). For MACE (Table S8), neither cohort showed significant associations across *APOE* genotypes, as shown in Panels C of Figures 3 and 4, with hazard ratios near 1 for all genotypes.

To evaluate the relationship between net changes in lipid biomarkers and all-cause mortality, we included each biomarker as an additional covariate in the analysis (Figure 5). The median net changes (in mmol/L) were as follows: for the UKB cohort, HDLC: 0.00, LDLC: -1.25, TC: -1.30, and TG: -0.15; and for the AoU cohort, HDLC: 0.03, non-HDL: -0.98, LDLC: -0.82, TC: -0.85, and TG: -0.10 (Table S3). In the UKB cohort, net changes in lipid biomarkers were not significantly associated with mortality. In contrast, in the AoU cohort, an increase in HDLC was strongly protective (HR: 0.26, 95% CI: 0.16–0.41 per mmol/L), while decreases in LDLC (HR: 0.82, 95% CI: 0.69–0.97 per mmol/L) and triglycerides (HR: 0.79, 95% CI: 0.72–0.87 per mmol/L) were significantly associated with a reduced risk of all-cause mortality.

**Figure 5.**
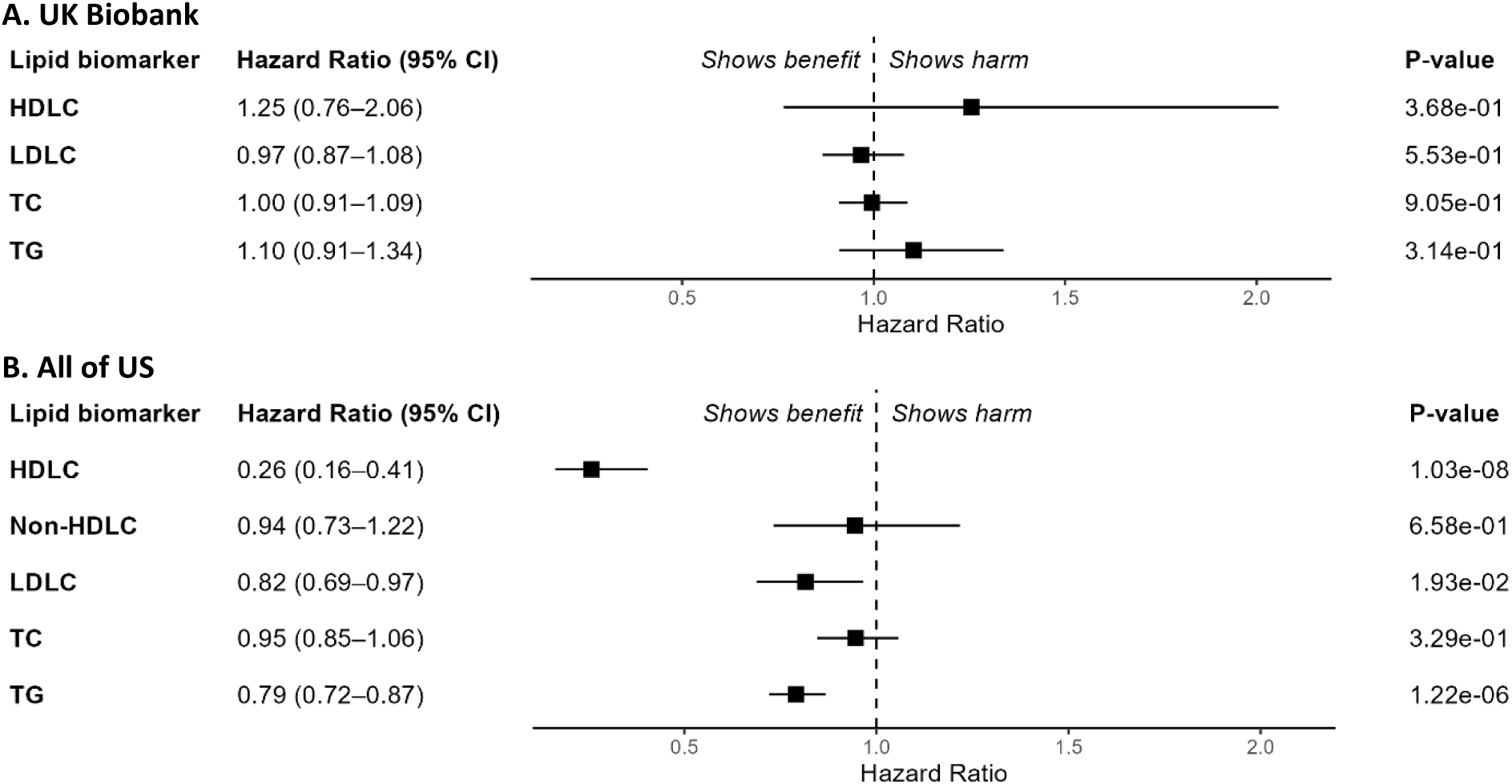
Associations between net changes (mmol/L) in lipid biomarkers and all-cause mortality. Panels **A** and **B** show hazard ratios (HRs) with 95% confidence intervals (CIs) for the UK Biobank and All of Us cohorts, respectively. Net changes in HDLC were calculated as post-treatment minus pre-treatment biomarker levels, meaning HRs represent the risk associated with a 1 mmol/L increase in HDLC (HR < 1 indicates a beneficial effect). For all other biomarkers, net changes were calculated as pre-treatment minus post-treatment levels, so HRs reflect the risk associated with a 1 mmol/L reduction (HR < 1 indicates a beneficial effect). Analyses were adjusted for age, sex, histories of hypertension, diabetes mellitus, and cardiovascular disease, data provider (UK Biobank), type and strength of statin, genotyping array (UK Biobank), *APOE* genotype, and the first ten principal components of genetic ancestry. *APOE* = Apolipoprotein E, HDLC, high-density lipoprotein cholesterol; LDLC, low-density lipoprotein cholesterol; TC, total cholesterol; TG, triglycerides.

## Discussion

Using electronic health records from the UK Biobank (UKB) and the All of Us (AoU) program, we examined the relationship between statin initiation and various clinical outcomes. Consistent with existing evidence, increases in HDLC and reductions in LDLC and triglycerides were protective in the AoU cohort in terms of the risk of all-cause mortality. In this cohort, the median net changes in lipid biomarkers (in mmol/L and percentage change) were as follows: HDLC: 0.03 (2%), LDLC: −0.82 (−27%), and triglycerides: −0.10 (−8.16%) while the UKB cohort changes were: HDLC: 0 (0%), LDLC: −1.25 (−35%), and triglycerides: −0.15 (−11.8%).

These changes are consistent with the expected benefits of statins, which inhibit HMG-CoA reductase, the key enzyme regulating cholesterol synthesis. Statins typically lead to a 1–10% increase in HDLC, a reduction of approximately 30% in LDLC for low-intensity statins, a 30–49% reduction in LDLC for moderate-intensity statins, and a 10–20% reduction in triglycerides.^11, 12, 38, 39^

Each 1 mmol/L increment or reduction in lipid biomarkers was associated with significant changes in mortality risk: HDLC (74% reduction in risk, HR: 0.26, 95% CI: 0.16–0.41), LDLC (18% reduction in risk, HR: 0.82, 95% CI: 0.69–0.97), and triglycerides (21% reduction in risk, HR: 0.79, 95% CI: 0.72–0.87). When combining the observed median changes in lipid biomarkers with these risk estimates, the corresponding statin-associated reductions in all-cause mortality in the AoU cohort were approximately 2.2% for HDLC (0.03 mmol/L increase × 74% reduction per mmol), 14.8% for LDLC, and 2.1% for triglycerides, indicating that the largest mortality benefit was associated with LDLC reduction. This mortality benefit per mmol/L is consistent with findings from a meta-analysis of 90,056 participants across 14 randomized controlled trials (RCTs), which reported a rate ratio of 0.88 (95% CI: 0.84–0.91) per mmol/L reduction in LDLC.^40^ The mean LDLC reduction at 1 year in these RCTs (1.09 mmol/L) is comparable to the reductions observed in our study (AoU: -0.82 mmol/L, UKB: -1.25 mmol/L). While biomarker changes were observed in both cohorts, associations with all-cause mortality were significant only in the AoU cohort. This discrepancy may be attributed to sample size differences: the AoU cohort included over 11,000 more participants in the HDLC analysis (17,626 vs. 5,736), over 6,000 more participants in the LDLC analysis (11,235 vs. 5,052), and over 12,000 more participants in the triglycerides analysis (17,875 vs. 10,977), potentially providing greater statistical power to detect significant associations.

Regarding the effect of *APOE* genotype, significant net or percentage changes were observed only for HDLC and triglycerides, while LDLC, the most impacted biomarker (in terms of effects on mortality), showed non-significant changes. However, a visual inspection of Figure 2 reveals a trend in percentage changes: *ε2ε3* carriers showed negative percentage changes, indicating greater LDLC reductions compared to *ε3ε3* carriers, whereas *ε3ε4* and *ε4ε4* carriers showed positive percentage changes, reflecting smaller LDLC reductions compared to *ε3ε3* carriers. This pattern aligns with previous findings that *ε2* carriers typically experience greater LDLC reductions compared to *ε3ε3* carriers, while *ε4* carriers tend to be less responsive to statins.^6–8^

The lack of statistical significance for LDLC changes is likely due to the relatively small sample size and even smaller genotype subgroups. To address this, a post-hoc analysis was performed by grouping *ε2ε2* (without dysbetalipoproteinemia) and *ε2ε3* carriers into an "*ε2* carrier" group and combining *ε3ε4* and *ε4ε4* carriers into an "*ε4* carrier" group. *ε2ε4* carriers were assigned to either the "*ε2* carrier" group (UKB) or the "*ε4* carrier" group (AoU), based on genotype-specific effect directions. This grouping improved statistical power, decreasing the LDLC p-value from 0.036 to 0.012 in the UKB cohort and from 0.223 to 0.030 in the AoU cohort. After regrouping, *ε2* carriers showed 2.8% greater reductions and *ε4* carriers showed 0.4% smaller reductions compared to *ε3* carriers in the UKB cohort. Similarly, in the AoU cohort, *ε2* carriers showed 1.7% greater reductions, while *ε4* carriers showed 0.7% smaller reductions relative to *ε3* carriers. These percentage changes reflect population averages – they were accompanied by wide confidence intervals like those in Figure 2 highlighting the small sample size relative to the high variability within the population. This variability suggests that these changes may be substantial for certain individuals, who could benefit from personalized medicine. In the post-hoc analysis, *ε2ε4* carriers were classified under both *ε2* and *ε4* carriers, a common practice.^10^ However, the literature lacks consensus on how to handle the *ε2ε4* genotype (whether to exclude it or classify it as *ε2*, *ε3*, or *ε4*) and grouping *ε2ε2* with *ε2ε3* may be misleading, given the former’s association with an increased risk of familial dysbetalipoproteinemia.^3, 9, 10^ These complexities informed our decision to focus on *APOE* genotypes rather than carrier status. Moreover, genotype-level effect estimates can be pooled to analyse carrier-level effects, but the reverse is not possible, highlighting the flexibility and interpretive advantages of genotype-specific analyses.

To maximize sample size, we analysed the association between *APOE* genotypes and clinical outcomes without adjusting for lipid biomarkers, as these data were available for only a small fraction of participants (one-seventh in UKB and half in AoU). Consistent with prior evidence linking the *ε4* allele to adverse cardiovascular and metabolic profiles,^6–8^ which may contribute to increased mortality, the *ε4* allele was significantly associated with all-cause mortality in the UKB cohort (N = 45,515). Specifically, *ε3ε4* carriers had a modestly elevated risk (HR: 1.08, 95% CI: 1.01–1.15), while *ε4ε4* carriers showed a substantially higher risk (HR: 1.54 [1.33–1.78] which slightly decreased to HR: 1.30 [1.01–1.68] for cardiovascular-related mortality). These findings are consistent with earlier analyses of the UK Biobank baseline assessment data and linked mortality records (452,189 participants), where *ε4ε4* carriers had a hazard ratio of 1.51 (95% CI: 1.41–1.62) for all-cause mortality and 1.54 (95% CI: 1.33–1.77) for cardiovascular-related mortality.^13^ In the smaller AoU cohort (N = 35,562), only *ε4ε4* carriers similarly showed a significantly increased risk of all-cause mortality (HR: 1.64, 1.08–2.49) with *ε3ε4* carriers showing a non-significant increase (HR: 1.07, 0.89–1.28). These findings are consistent with our power analyses (Figure S2), which indicated greater power to detect associations in the larger UKB cohort compared to the AoU cohort.

Despite several strengths, including two relatively large cohorts (one homogeneous and one diverse) and robust associations supported by sensitivity analyses (e.g., excluding dysbetalipoproteinemia participants and conducting complete-case analysis for AoU), our study had limitations. These included low statistical power for some outcomes and the inability to incorporate relevant covariates such as body mass index or adherence to statin treatment, either due to challenges in accurate phenotyping or the absence of these variables in the electronic health records used.^41^ Additionally, the representativeness of the cohorts should be considered; for example, UKB participants have been reported to be, on average, healthier than the general population.^42^

In conclusion, our analysis highlights the impact of *APOE* genotype on clinical outcomes, particularly all-cause mortality. *ε4* carriers, especially *ε4ε4*, demonstrated significantly increased mortality risks in both the UKB and AoU cohorts. Consistent with prior evidence, statin-induced increases in HDLC and reductions in LDLC and triglycerides were associated with lower all-cause mortality. Larger studies are needed to further strengthen the evidence base and explore associations with other clinical outcomes that could not be detected due to limited statistical power. Our results emphasize the importance of exploring the interplay between *APOE* genotype, lipid profiles, and clinical outcomes, particularly in diverse populations and varying cohort contexts. They also reaffirm that *APOE* genotype significantly influences statin response, emphasizing the value of incorporating genetic information into personalized treatment strategies.

## Conflict of interest

M.P. currently receives partnership funding, paid to the University of Liverpool, for the following: MRC Clinical Pharmacology Training Scheme (co-funded by MRC and Roche, UCB, Eli Lilly and Novartis), and the MRC Medicines Development Fellowship Scheme (co-funded by MRC and GSK, AZ, Optum and Hammersmith Medicines Research). He has developed an HLA genotyping panel with MC Diagnostics but does not benefit financially from this. He is part of the IMI Consortium ARDAT (www.ardat.org); none of these of funding sources have been used for the current research. All other authors declared no competing interests for this work.

## Funding

This work was supported by the Medical Research Council [MR/V033867/1; Multimorbidity Mechanism and Therapeutics Research Collaborative].

## Data Availability

The data that support the findings of this study are available from UK Biobank (https://www.ukbiobank.ac.uk/) and All of Us Research Program (https://allofus.nih.gov/), with the permission of UK Biobank and All of Us Research Program, respectively.

## Notes

### Multimorbidity Mechanism and Therapeutic Research Collaborative

Adam Butterworth, Alasdair Warwick, Alba Fernandez-Sanles, Albert Henry, Alvina G Lai, Amanda Roberts, Andrea Jorgensen, Ana Torralbo, Anoop D Shah, Aroon Hingorani, Arturo Gonzalez-Izquierdo, Maria Carolina Borges, Caroline Dale, Chris Finan, Claudia Langenberg, Daniel C Alexander, Deborah Lawlor, Diana Dunca, Eda B Ozyigit, Amand F Schmidt, Harry Hemingway, Honghan Wu, Innocent G Asiimwe, Jasmine Gratton, Jean Gallagher, Jorgen E Engmann, Lauren E Walker, Victoria L Wright, Magdalena Zwierzyna, Margaret Ogden, Martin Cox, Mary Mancini, Michail Katsoulis, Mira Hidajat, Munir Pirmohamed, Natalie Fitzpatrick, Nishi Chaturvedi, Rashmi Kumar, Rohan Takhar, Sandesh Chopade, Simon Ball, Spiros Denaxas, Tina Shah, Valerie Kuan, Nikita Hukerikar, Reecha Sofat, Frances Bennett, David Ryan, Maik Pietzner.

## Acknowledgements

This research has been conducted using the UK Biobank Resource under Application Number 56653. We gratefully acknowledge All of Us participants for their contributions, without whom this research would not have been possible. We also thank the National Institutes of Health’s All of Us Research Program for making available the participant data examined in this study.

## Notes

### Author Declarations

Ethical approval was received by the UK Biobank (North-West Multicentre Research Ethics Committee approval number: 11/NW/0382), and written informed consent was received from all participants. Our study was approved by the UK Biobank (application number: 56653). The All of Us Research Program dataset v7 repository includes electronic health records (EHR) and genomic data, accessible through a cloud-based Researcher Workbench. Before the program could begin to recruit and enroll participants, the All of Us IRB had to approve the All of Us protocol and materials (https://allofus.nih.gov/about/who-we-are/institutional-review-board-irb-of-all-of-us-research-program). Access to genomic data requires Controlled tier access, granted after completing registration, training, and a data-use agreement.

