## Supplementary Figures for "*APOE* Genotype and Statin Response: Evidence from Electronic Health Records in the UK Biobank and All of Us Research Program"

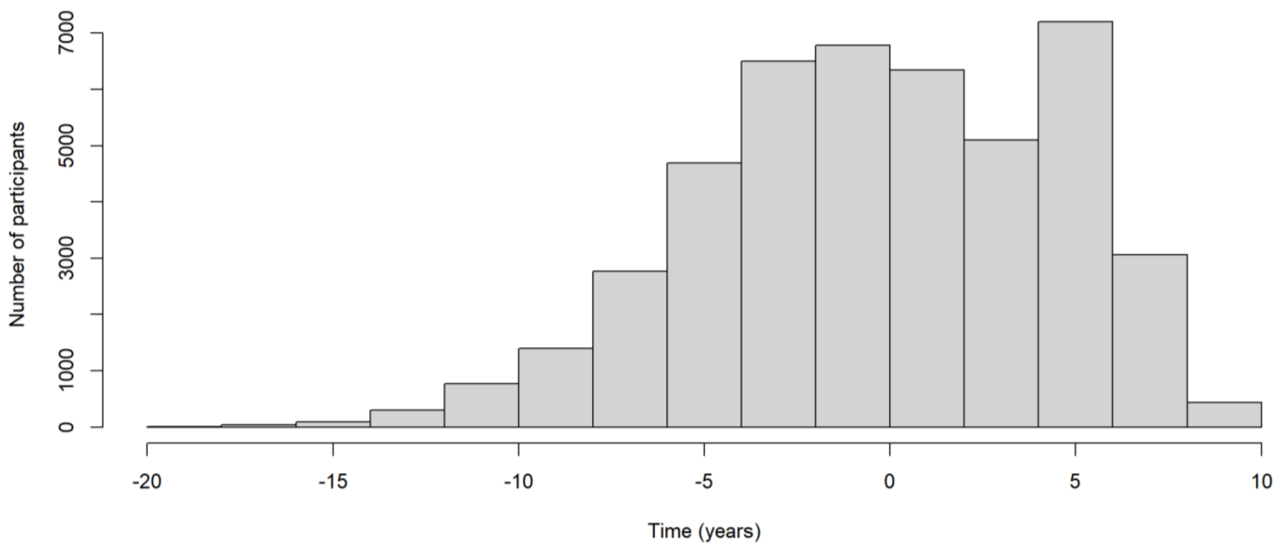


**Figure S1. Time difference between the index date (start of follow-up) and the date of attending a UK Biobank assessment centre.**

### A. UK Biobank

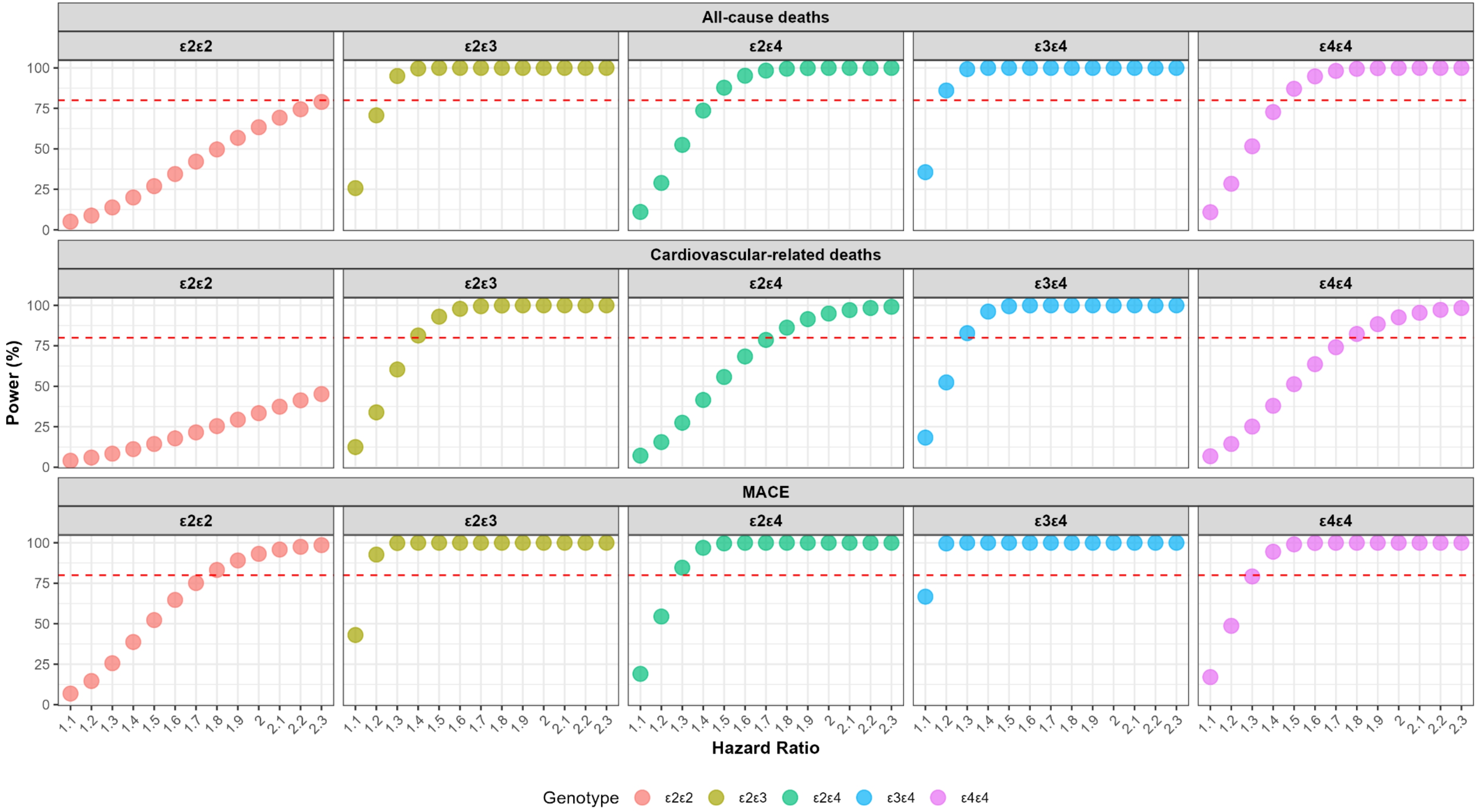

All-cause deaths
Cardiovascular-related deaths
MACE
ε2ε2
ε2ε3
ε2ε4
ε3ε4
ε4ε4
Power (%)
0
25
50
75
100
1.1
1.2
1.3
1.4
1.5
1.6
1.7
1.8
1.9
2
2.1
2.2
2.3
Hazard Ratio
Genotype
ε2ε2
ε2ε3
ε2ε4
ε3ε4
ε4ε4

**B. All of Us**

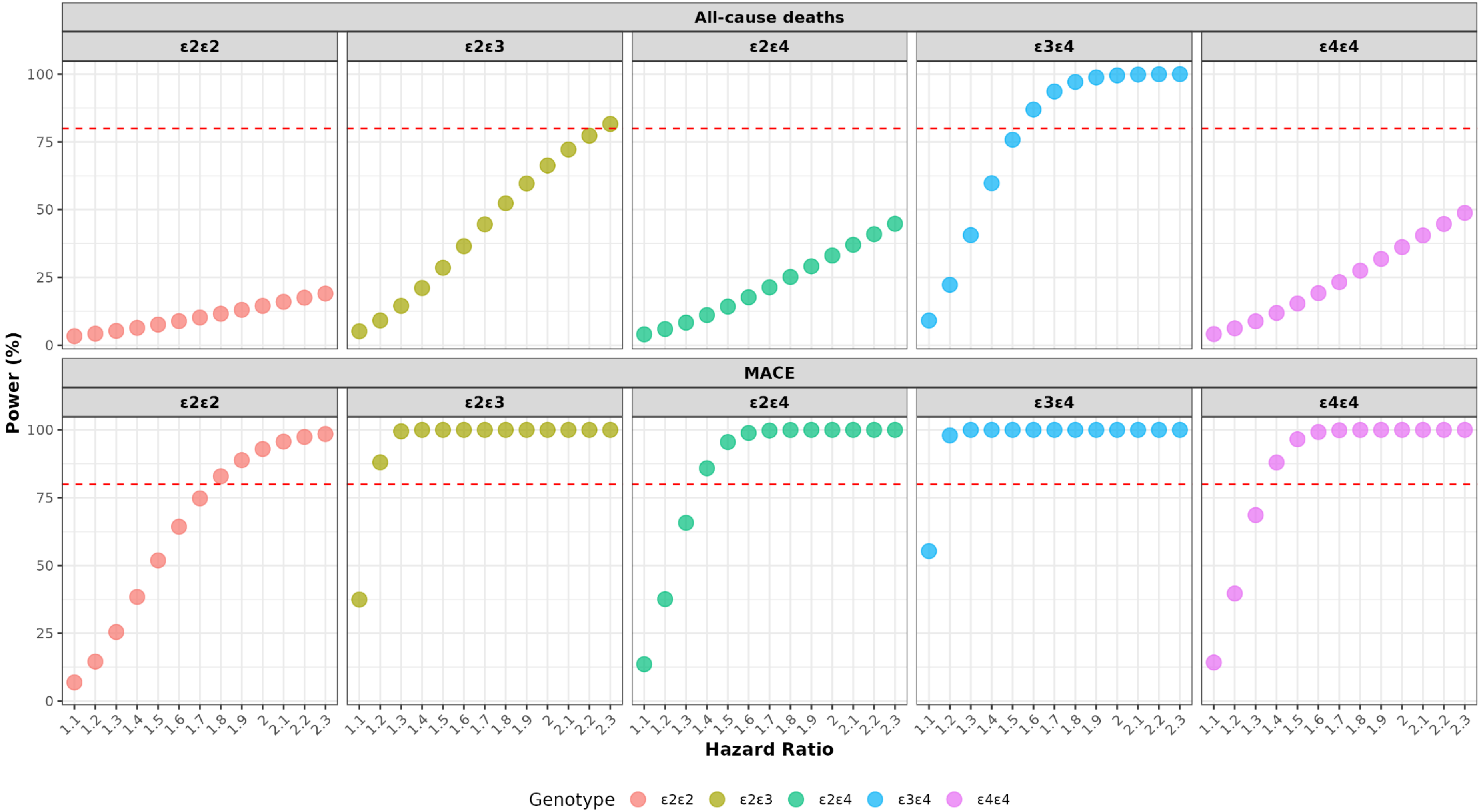


**Figure S2. Power analysis for varying hazard ratios for each *APOE* genotype, with *ε3ε3* as the reference genotype. A. UK Biobank.** Sample sizes of 45,515 and 45,149 were assumed for mortality and major adverse cardiovascular events (MACE) outcomes, respectively. **B. All of Us Program.** A sample size of 35,562 was assumed. Panels display power (%) to detect hazard ratios across the genotypes *ε2ε2*, *ε2ε3*, *ε2ε4*, *ε3ε4*, and *ε4ε4*. The dashed red line indicates the 80% power threshold.

**A. Convergence Plot**

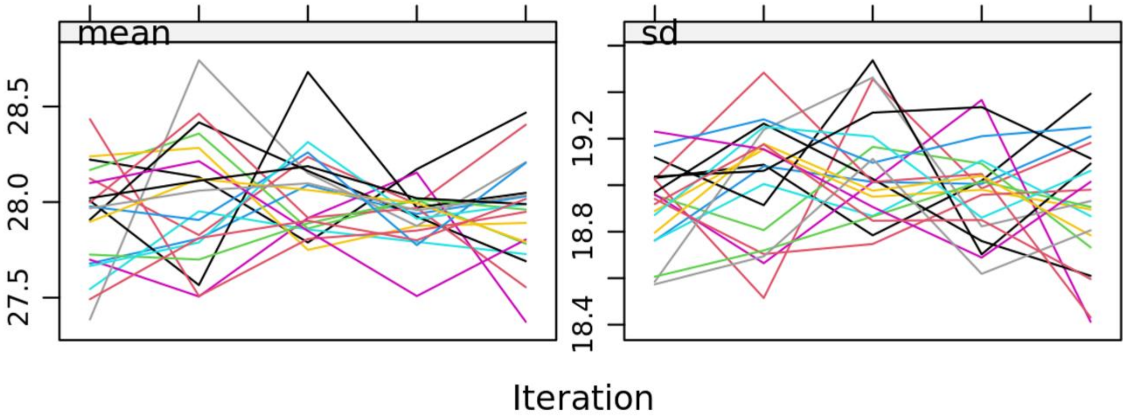


**B. Density plot**

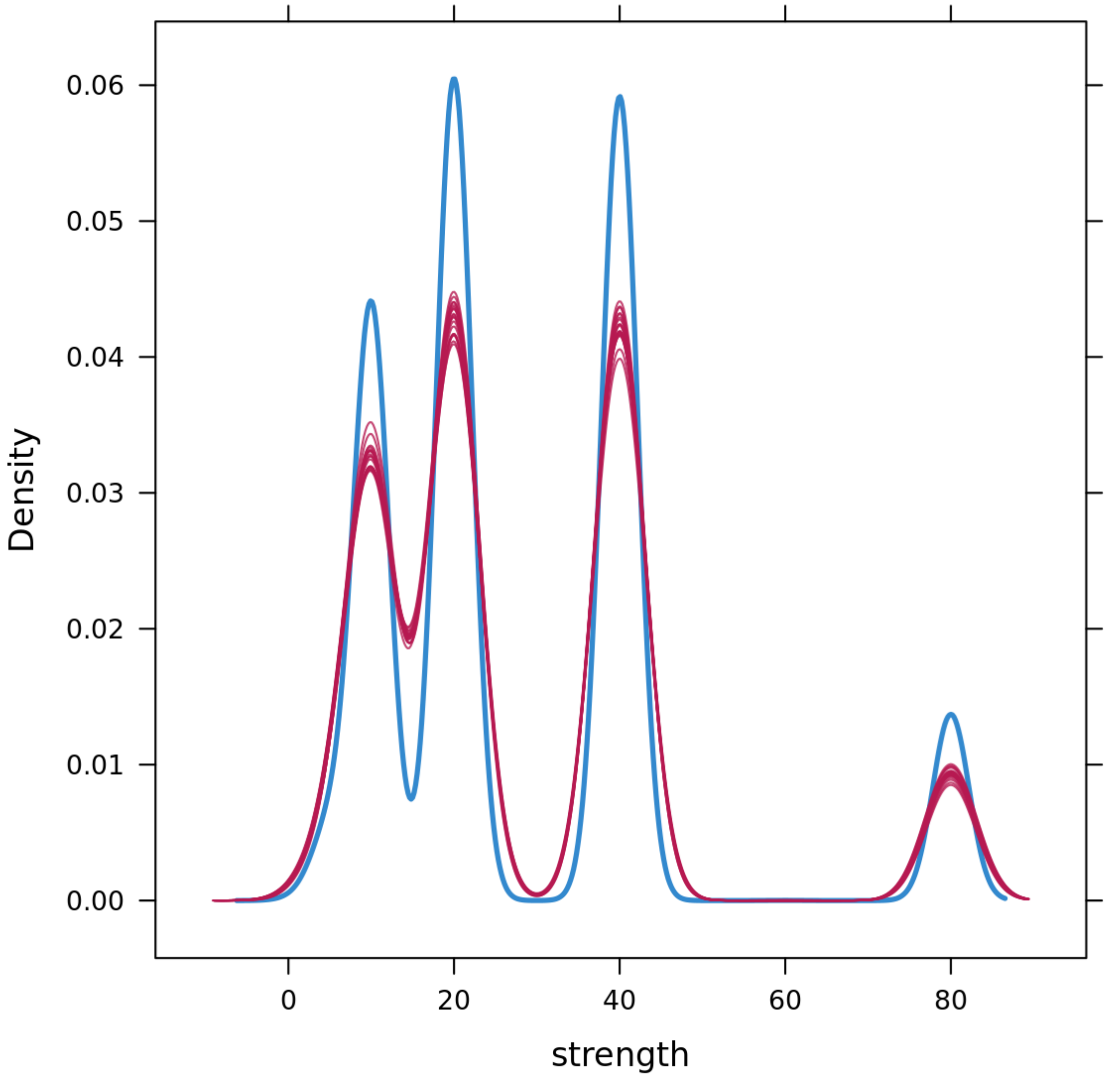


**Figure S3. Imputation assessment for statin strength.** The convergence plot (**Panel A**) shows stable mean and standard deviation values across iterations, indicating that the imputation algorithm reached convergence. Each line represents a different chain, with mean values stabilizing around 27.5–28.5 and standard deviation (sd) around 18.4–19.5. The density plot (**Panel B**) displays overlapping density curves from various iterations (blue: observed; red: imputed) showing the consistency of imputed statin strength values.

**A. UK Biobank (pre-statin measurements)**

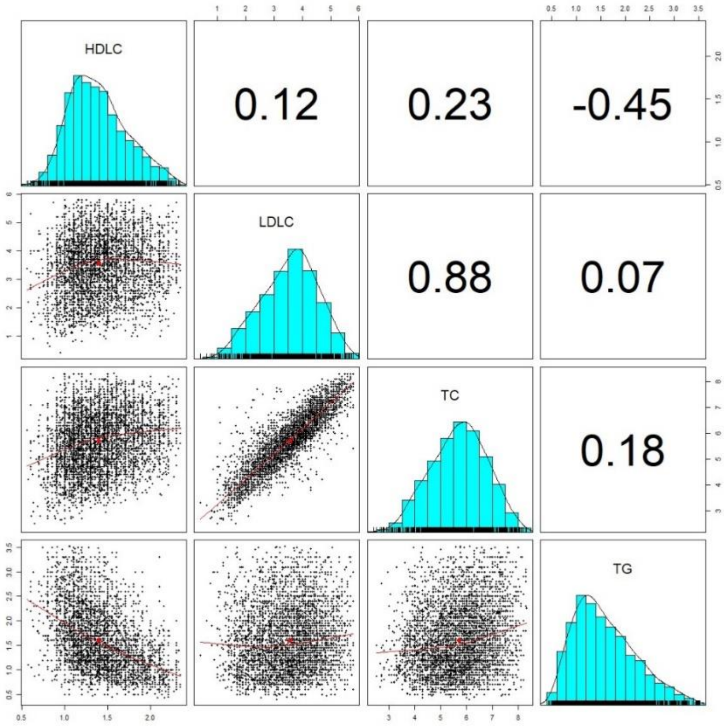


**B. UK Biobank (net changes)**

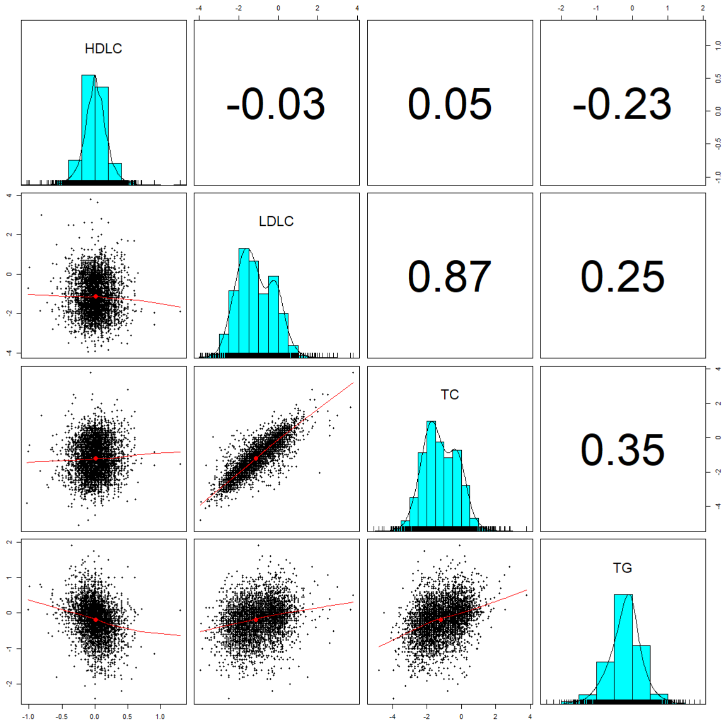

**C. All of Us (pre-statin measurements)**

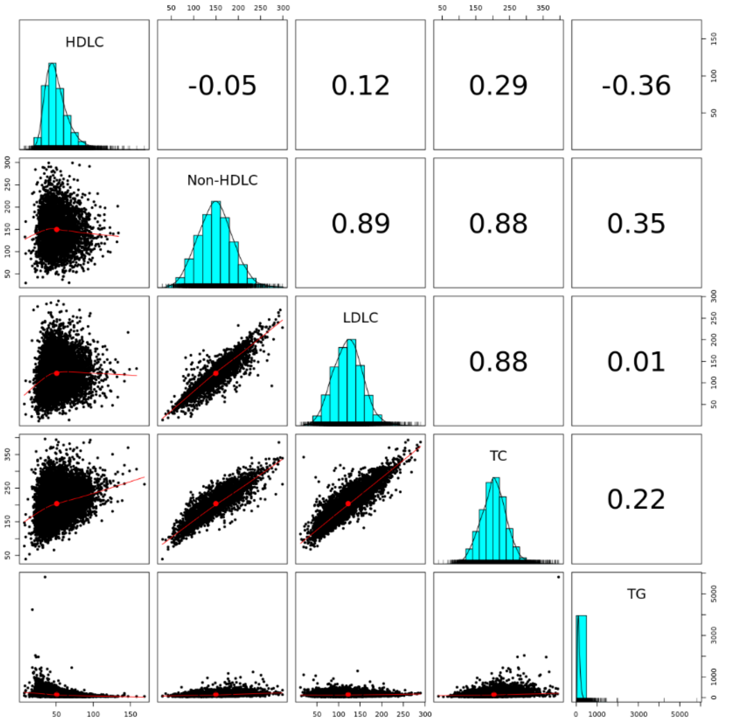


**D. All of Us (net changes)**

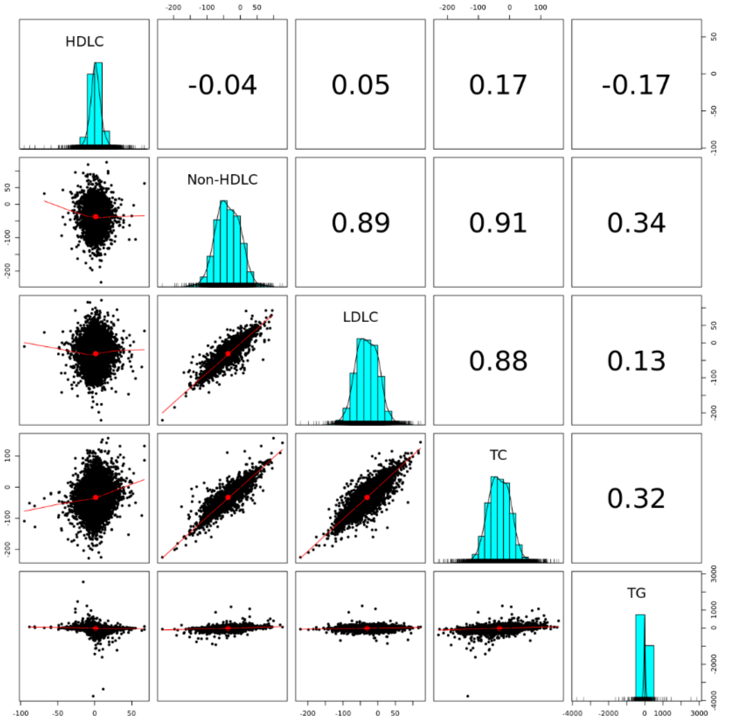


**Figure S4. An enhanced scatterplot matrix showing correlations between the lipid biomarkers. A.** UK Biobank pre-statin measurements. **B.** UK Biobank net changes. **C.** All of Us pre-statin measurements. **D.** All of Us net changes. This figure shows a correlation matrix and pairwise scatter plots for key lipid biomarkers: HDL cholesterol (HDLC), LDL cholesterol (LDLC), non-HDLC, total cholesterol (TC), and triglycerides (TG). The upper triangles show Pearson correlation coefficients between the biomarkers, the diagonals display the distribution of each biomarker, and the lower triangles shows scatter plots with locally weighted smoothing (red lines) to depict relationships between the variables.

**A. UK Biobank**

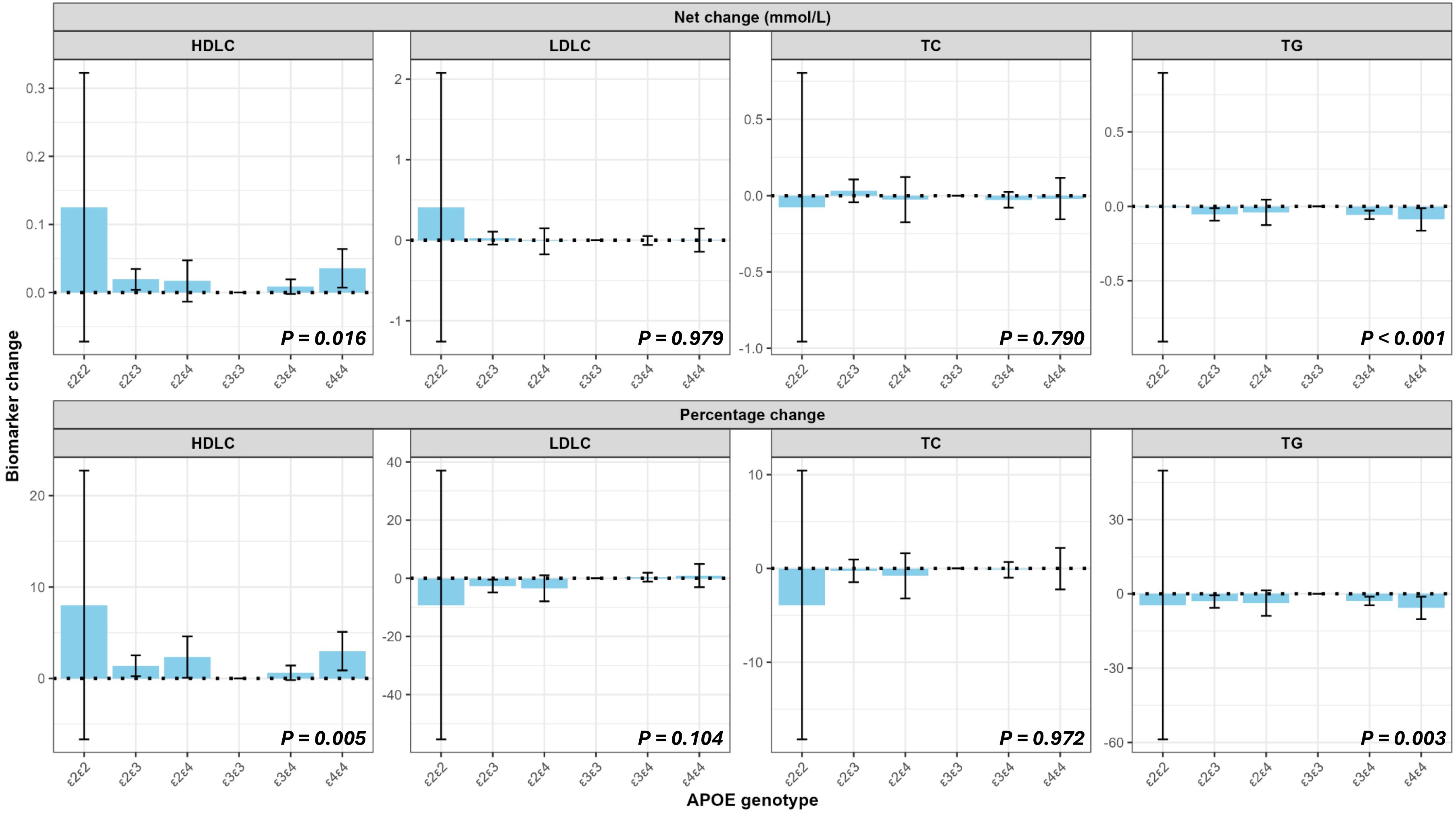

Net change (mmol/L)
HDLC
LDLC
TC
TG
P = 0.016
P = 0.979
P = 0.790
P < 0.001
Percentage change
P = 0.005
P = 0.104
P = 0.972
P = 0.003
Biomarker change
APOE genotype
ε2ε2
ε2ε3
ε2ε4
ε3ε3
ε3ε4
ε4ε4

**B. All of Us**

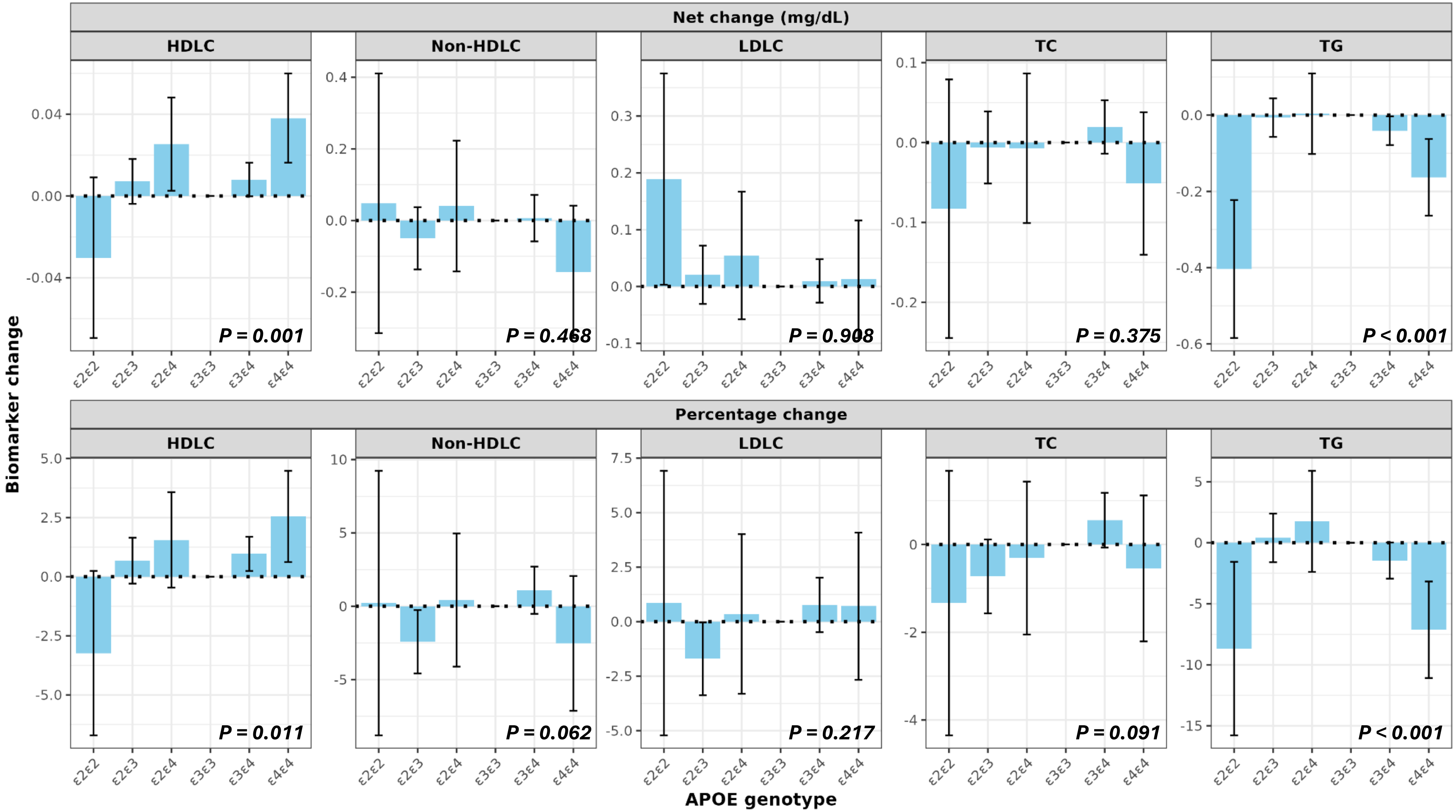


**Figure S5. Net and percentage changes in lipid biomarkers stratified by *APOE* genotype with dysbetalipoproteinemia individuals excluded. A.** UK Biobank. **B.** All of Us Program. The top and bottom rows respectively show net and percentage changes. For HDLC (increase beneficial), positive values indicate more benefit with statins relative to *ε3ε3*, while for other biomarkers (reduction beneficial), negative values indicate more benefit. Error bars represent 95% confidence intervals. *APOE* = Apolipoprotein E, HDLC = high-density lipid cholesterol, LDLC = low-density lipid cholesterol, TC = Total cholesterol, TG = triglycerides.

**A. UK Biobank**

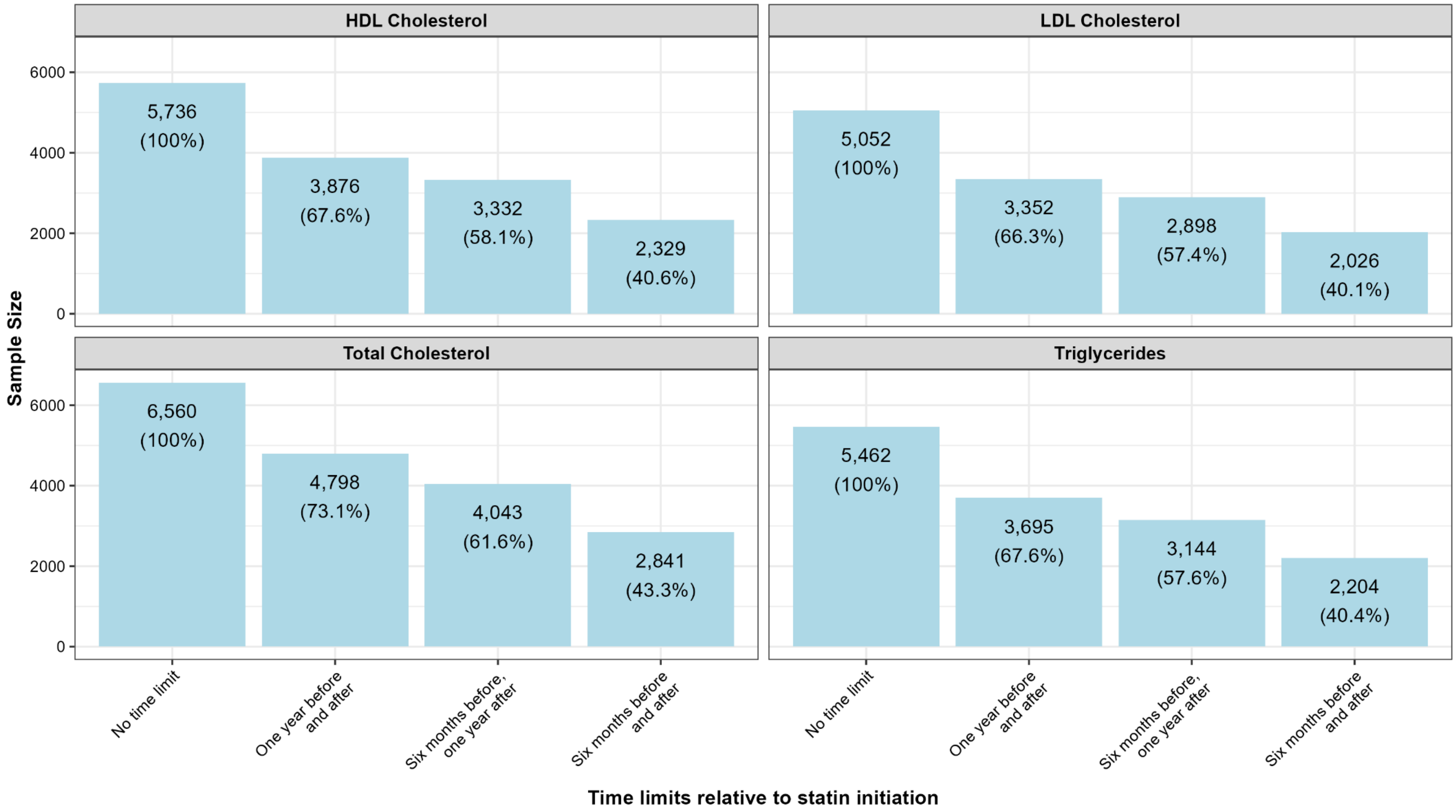
HDL Cholesterol
LDL Cholesterol
Total Cholesterol
Triglycerides
Sample Size
6000
4000
2000
0
5,736 (100%)
3,876 (67.6%)
3,332 (58.1%)
2,329 (40.6%)
5,052 (100%)
3,352 (66.3%)
2,898 (57.4%)
2,026 (40.1%)
6,560 (100%)
4,798 (73.1%)
4,043 (61.6%)
2,841 (43.3%)
5,462 (100%)
3,695 (67.6%)
3,144 (57.6%)
2,204 (40.4%)
No time limit
One year before and after
Six months before, one year after
Six months before and after
Time limits relative to statin initiation

**B. All of Us program**

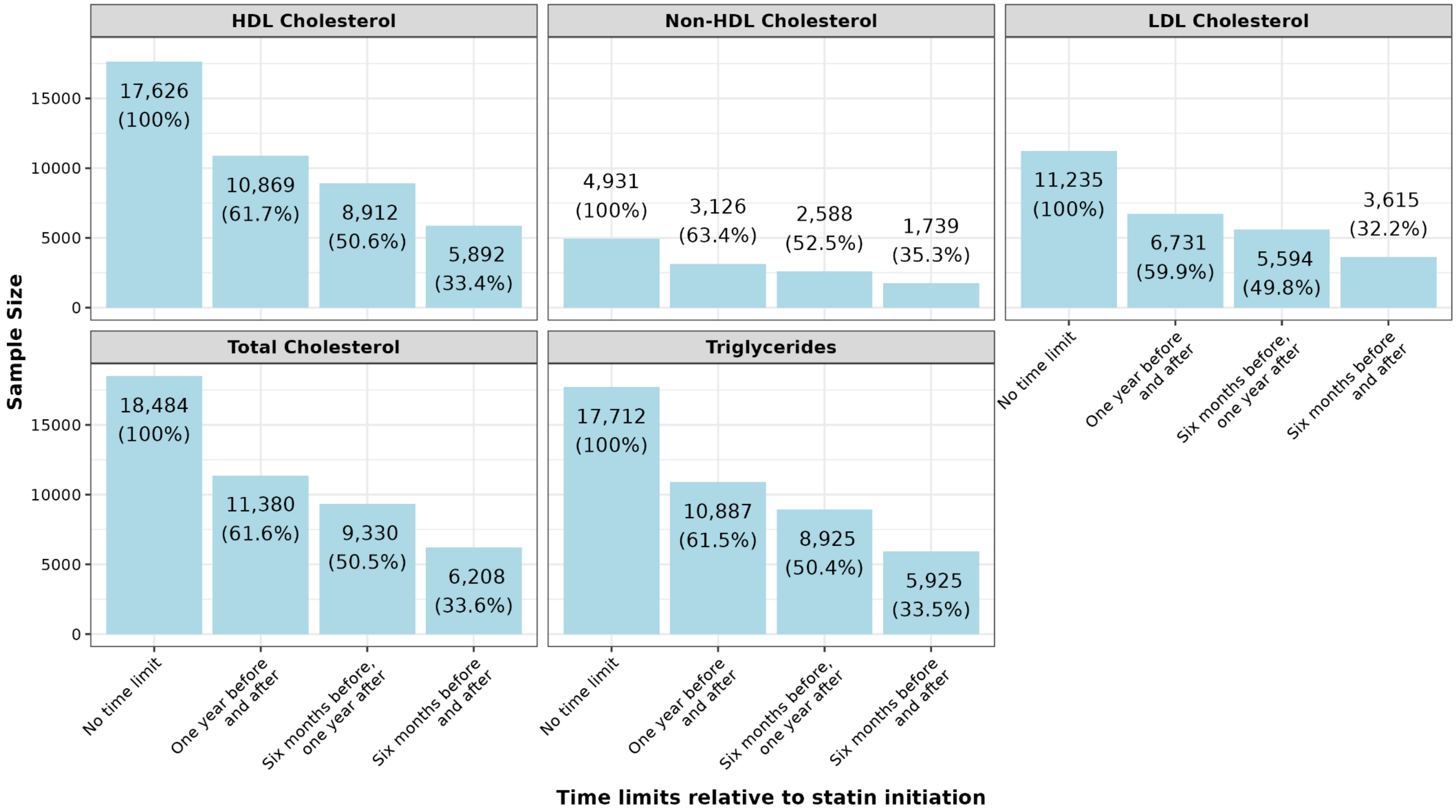


**Figure S6. Sample size across different time limits. A.** UK Biobank. **B.** All of Us program.  HDL = high-density lipoprotein, LDL = low-density lipoprotein.

### A. UK Biobank

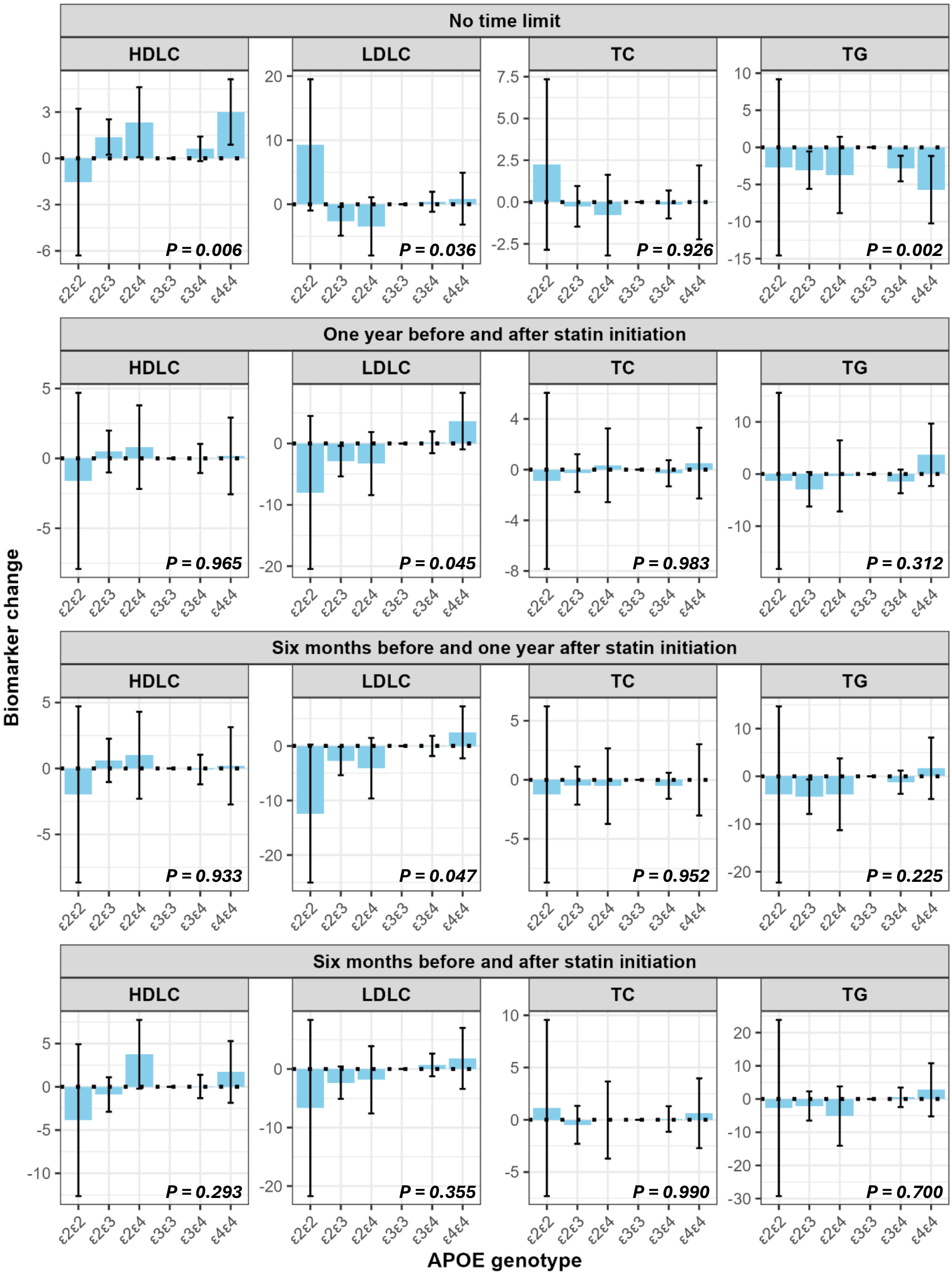

No time limit
One year before and after statin initiation
Six months before and one year after statin initiation
Six months before and after statin initiation
HDLC
LDLC
TC
TG
P = 0.006
P = 0.036
P = 0.926
P = 0.002
P = 0.965
P = 0.045
P = 0.983
P = 0.312
P = 0.933
P = 0.047
P = 0.952
P = 0.225
P = 0.293
P = 0.355
P = 0.990
P = 0.700
ε2ε2
ε2ε3
ε2ε4
ε3ε3
ε3ε4
ε4ε4
Biomarker change
APOE genotype

**B. All of Us**

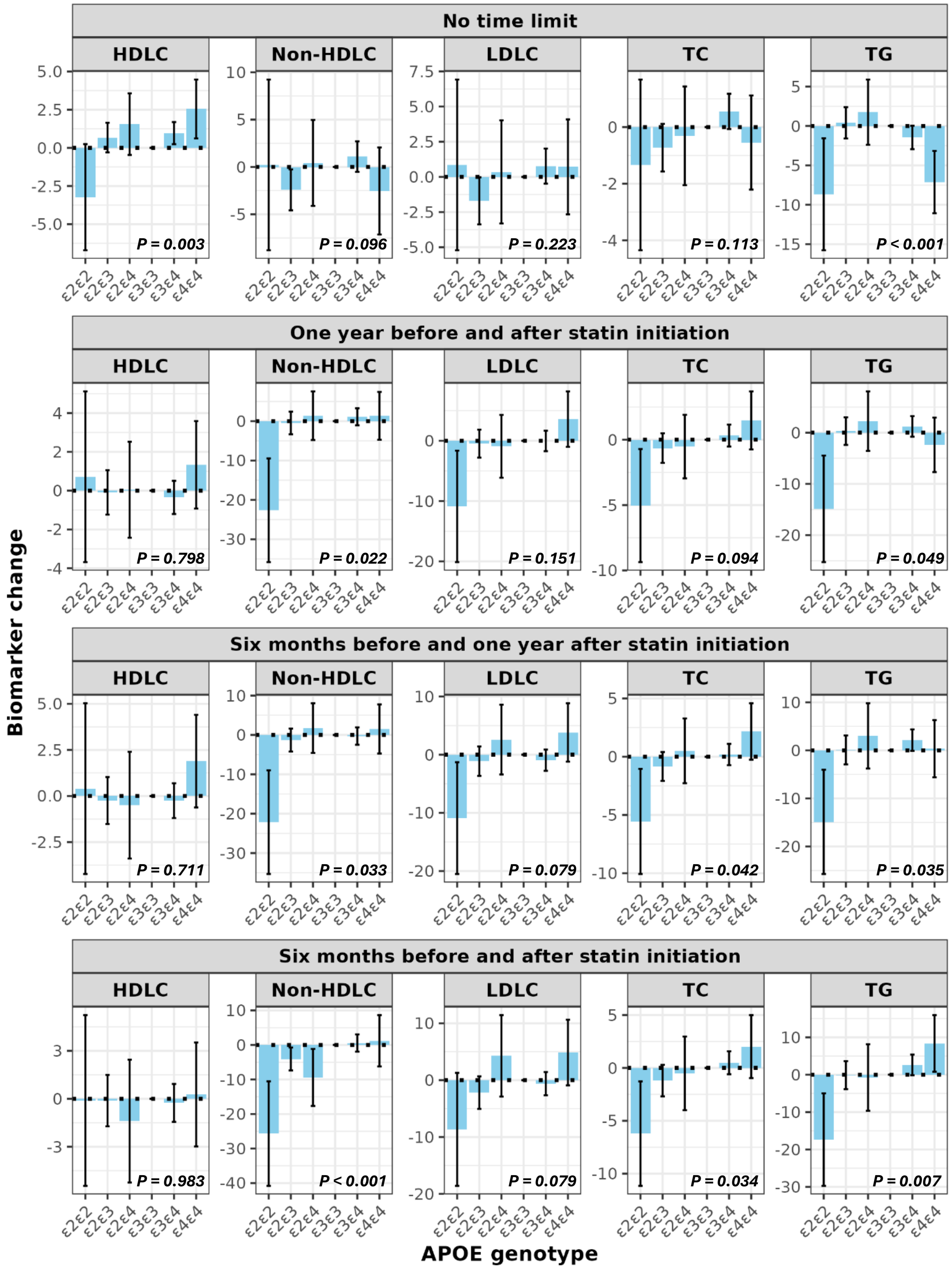


**Figure S7. Percentage changes in lipid biomarkers stratified by *APOE* genotype and the different time limits. A.** UK Biobank. **B.** All of Us Program. For HDLC (increase beneficial), positive values indicate more benefit with statins relative to the *ε3ε3* genotype, while for other biomarkers (reduction beneficial), negative values indicate more benefit. Error bars represent 95% confidence intervals. *APOE* = Apolipoprotein E, HDLC = high-density lipid cholesterol, LDLC = low-density lipid cholesterol, TC = Total cholesterol, TG = triglycerides.
